# Enhancing stress regulation in ecologically valid contexts through functional near-infrared spectroscopy neurofeedback of the prefrontal cortex

**DOI:** 10.1101/2025.07.29.25332351

**Authors:** Michelle Hei Lam Tsang, Judy Chuyi Chen, Heng Jiang, Benjamin Becker

## Abstract

**Significance:** Stress represents a key contributor to internalizing disorders and of rising global mental health challenges, particularly in young individuals. There is a critical need for accessible, brain-based interventions that can strengthen stress regulation and promote resilience.

**Aim:** We investigated whether a single session of real-time functional near-infrared spectroscopy (fNIRS)-informed neurofeedback training targeting the lateral prefrontal cortex (PFC) can enhance stress regulation under ecologically valid physiological and social stress conditions.

**Approach:** In a pre-registered double-blind, sham-controlled parallel-group trial, 60 young healthy adults underwent four neurofeedback, preceded and followed by baseline and maintenance runs without feedback. The training combined continuous feedback from individualized lateral PFC channels with reappraisal strategies to guide regulatory control. Stress regulation was assessed using the socially-evaluated cold pressor test (SECPT) that combines physiological and social stressors.

**Results:** Neurofeedback significantly increased lateral PFC activity across training runs. Participants in the neurofeedback group exhibited reduced stress – but not pain – experience in the SECPT and post-training anxiety, reflecting a successful domain-specific transfer of regulatory control.

**Conclusions:** This study demonstrates fNIRS-guided PFC neurofeedback facilitates adaptive learning of regulatory control and robustly enhances stress regulation and resilience under real-world stress. This scalable, non-invasive intervention offers a promising translational strategy for promoting resilience in vulnerable populations.

## 1 Introduction

Stress represents a transdiagnostic factor underlying the development, maintenance, and treatment of diverse mental health disorders, including internalizing disorders. The global prevalence of these disorders, in particular major depressive disorder and anxiety disorders, remains a rising trend following recent global crisis events (COVID-19 Mental Disorders Collaborators, 2021), with particularly pronounced increases among children, adolescents, and young adults (Kieling et al., 2024; Liu et al., 2025). This trend contrasts sharply with the lack of novel and efficacious interventions for at-risk youth, a disparity that is particularly concerning given that most mental health disorders emerge during adolescence (Solmi et al., 2022; Kessler et al., 2005), and lead to sustained psychological distress and functional impairments across the lifespan (Uhlhaas et al., 2023; Caspi et al., 2014). Given the mounting evidence of the personal and societal burden imposed by these conditions, developing innovative approaches to stress regulation emerges as a critical priority to foster resilience among vulnerable populations.

Neurofeedback is an effective neuromodulation technique based on biofeedback principles, where individuals receive real-time information on their physiological activity to guide self-regulation (Sitaram et al., 2017). Neurofeedback training represents a closed-loop approach during which brain activity is recorded and processed in (near) real-time, and indices of regional neural activity or functional connectivity are presented to participants, enabling direct engagement with neural substrates underlying cognitive and emotional processes (Gaume et al., 2016). Through this iterative learning process, individuals develop enhanced self-regulation abilities that improve behavioral and adaptive functioning (Paret et al., 2019).

The field of real-time neurofeedback, initially advanced through functional magnetic resonance imaging (fMRI), has risen in the past two decades as a pioneering approach for novel brain-based interventions in clinical populations (Linhartova et al., 2019). These interventions strategically target the well-established neural circuitry underpinning emotional processing (Etkin et al., 2015; Ochsner et al., 2012): limbic regions involved in fear and anxiety, and prefrontal regions that exert top-down cognitive control to downregulate these stress responses (Zimmermann et al., 2017; Zhuang et al., 2021; Mihov et al., 2013).

Previous fMRI neurofeedback studies have successfully capitalized on this framework, and have for example trained participants to volitionally regulate regional amygdala (e.g., Zotev et al., 2011; Young et al., 2014) and insula (e.g., Yao et al., 2016; Zhang et al., 2023), with recent studies directly targeting the dynamic interplay between prefrontal cortex (PFC) and the amygdala (e.g., Zhao et al., 2019; Paret et al., 2016). Successful control over brain activity was accompanied by region-specific changes in affective domains, including amygdala training reducing anxiety symptoms (Young et al., 2014; Zhao et al., 2019), and insula training improving empathic responses (Yao et al., 2016) and interoceptive awareness (Zhang et al., 2023). Notably, these improvements were maintained in the absence of feedback and maintained for several days after the training (e.g. Yao et al., 2016).

Collectively, these studies successfully demonstrate that neurofeedback training produces beneficial and sustained changes in affect regulation and behavioral outcomes among both healthy populations and clinical samples. Nevertheless, several limitations impede the scalability of fMRI approaches for real-life applications: methodological constraints include limited temporal resolution and sensitivity to motion artifacts, while implementation barriers encompass considerable operational costs and a confined scanning environment that limits its practicality for delivering multiple sessions typical of interventions.

Functional near-infrared spectroscopy (fNIRS) is a promising avenue for neurofeedback (Kohl et al., 2020; Soekadar et al., 2021), addressing several limitations of fMRI applications. Through the transmission and detection of near-infrared light in biological tissue, fNIRS measures hemoglobin concentrations to infer regional activity based on neurovascular coupling principles, similar to the blood-oxygenation level-dependent (BOLD) signal in fMRI (Steinbrink et al., 2006). fNIRS offers methodological capabilities in capturing cortical hemodynamic activity with considerable temporal precision, combined with motion tolerance and implementation in naturalistic environments. These advantages position fNIRS as a distinct neuroimaging modality that bridges the gap between laboratory research and practical applications, serving as an ideal platform for neurofeedback interventions that train regulatory skills in real-world contexts where they are most critically needed.

As a relatively young, yet rapidly expanding field, fNIRS neurofeedback has been applied to strengthen various cognitive functions, with marked success in targeting the lateral PFC due to its key involvement in several important domains (Friedman & Robbins, 2022; Ray & Zald, 2012). Existing studies have demonstrated success in improving working memory (Li et al., 2023; Yang et al., 2024; Hou et al., 2021), cognitive flexibility (Li et al., 2019) and conflict adaptation (Tang et al., 2021) through modulating the lateral PFC. Moreover, studies utilizing lateral PFC neurofeedback with general cognitive strategies have shown improvements in subsequent emotion regulation tasks, highlighting this region’s capacity in broadly training regulatory functions (Yu et al., 2021; Li et al., 2025). Given the consistent meta-analytic identification of the lateral PFC as a critical node in emotion regulation (Kohn et al., 2014; Picó-Pérez et al., 2017; Zilverstand et al., 2017), there is a compelling rationale for fNIRS neurofeedback applications targeting the lateral PFC (for enhancing stress regulation). Several approaches have shown promise within the emotion processing domain through neurofeedback interfaces to modulate affective processes (Aranyi et al., 2016; Trambaiolli et al., 2018) and attenuate threat processing (Kimmig et al., 2019). Building on these advances, recent studies (Yang et al., 2024; Godet et al. 2024) have shown that lateral PFC training elicits changes in connectivity extending to subcortical regions, indicating that strategically targeting this region could potentially modulate emotional reactivity at limbic centers via regulatory neural pathways.

Lateral PFC neurofeedback via fMRI has been paired with explicit training instructions utilizing cognitive reappraisal, a well-validated emotion regulation strategy involving the reinterpretation of aversive stimuli to diminish their negative emotional impact (Gross, 2002). The usage of reappraisal strategies strengthens functional connectivity between lateral PFC and the amygdala, facilitating a reliable increase of PFC activation concurrent with decreased amygdala activity (Berboth & Morawetz, 2021; Buhle et al., 2014). Initial studies have shown that integrating cognitive reappraisal training with lateral PFC neurofeedback produces positive benefits among clinical populations, reducing negative affect and symptomatology in depression (Keller et al., 2021), anxiety (Zilverstand et al., 2015), and trauma-related disorders (Zweerings et al., 2020). Thus, neurofeedback targeting lateral prefrontal regions in combination with cognitive reappraisal strategies represents a precision-guided approach that strengthens voluntary control over prefrontal regulatory processes.

Despite growing research into neurofeedback applications, several key considerations are necessary to facilitate its translation into practical stress regulation interventions. First, precise targeting that matches brain regions with complementary regulation strategies is essential for consistent neurofeedback effects (Stoeckel et al., 2014) and successful regulation outcomes (Linhartová et al., 2019). Cognitive reappraisal as an explicit regulation strategy provides clear mapping to lateral PFC (Zilverstand et al., 2017; Pico-Perez et al., 2017), yet remains a minority approach in neurofeedback protocols despite numerous studies demonstrating its efficacy in producing lasting, transferable benefits within clinical populations (Zilverstand et al., 2015; Zweerings et al., 2020; Keller et al., 2021). Moreover, several existing studies in this domain lack appropriate control groups and adequate sample sizes, making it difficult to delineate neurofeedback-specific effects from confounds such as general engagement, use of reappraisal strategies, or placebo effects (Webb et al., 2012; Thibault et al., 2018). Lastly, it remains critical to establish whether neurofeedback training enhances regulatory capacity in real-world contexts, yet many studies rely predominantly on self-report measures, which reportedly produce small effect sizes particularly on affective tasks (Chiasson et al., 2023). Assessment of training outcomes necessitates empirically testing generalization of regulatory skills beyond training settings, particularly achievable via functionally-relevant standardized tasks that replicate naturalistic stress, in order to inform the translation of fNIRS neurofeedback into accessible interventions for at-risk populations (Krause et al., 2024).

The present investigation examines the efficacy and translational utility of real-time fNIRS neurofeedback targeting lateral prefrontal regions in conjunction with cognitive reappraisal strategies for stress regulation in healthy young adults, advancing translational applications by leveraging fNIRS technology through an optimized neurofeedback protocol and evaluating stress reactivity under naturalistic conditions in the absence of feedback. Using a pre-registered, randomized double-blind sham-controlled single-session protocol, 60 participants were randomly allocated to either real-time neurofeedback (experimental group) or yoked feedback (control group). This controlled approach is essential to determine whether the combination of fNIRS training of the lateral PFC enhances the effects of reappraisal as an emotion regulation strategy and leads to regulatory control over this brain region. The socially-evaluated cold pressor test (SECPT), a standardized stress induction paradigm combining physiological and social stressors, was implemented after the training to examine whether the neurofeedback training translates into enhanced stress regulation, with pain ratings serving as a contrast to assess domain-specificity of regulatory effects.

Our investigation established the following outcomes: The primary outcome measure assessed activity in trained target regions measured via fNIRS, with analyses determining whether neurofeedback increases activity in individualized target channels across the training. Secondary outcomes were implemented to evaluate whether neurofeedback enhances regulation beyond the training, operationalized through subjective reports of stress and pain from the SECPT. Additional analyses examined behavioral outcomes, including negativity ratings in response to images during the training and self-report ratings of anxiety, valence and arousal, as well as the regional specificity of neurofeedback training by analyzing channel activity patterns across the runs. The methodological rigor of our design facilitates examination of whether targeting emotion regulation circuitry yields specific neural modulation effects that extend beyond placebo responses or reappraisal practice alone, while validating the application of neurofeedback for stress regulation in individuals.

## 2 Materials and Methods

### 2.1 Experimental Design

#### 2.1.1 Participants

This study recruited 60 healthy young adults (18-27 years) from the Asian student population at the University of Hong Kong. Given limited power analysis standards for both fNIRS and neurofeedback studies, this sample size was determined based on previous studies in the field (see Li et al., 2023; Yang et al., 2024).

Participants were recruited through the university participant pool and email advertisements, with compensation provided through course credit or monetary reimbursement, respectively. Eligible participants were recruited if they met the following inclusion criteria: 1) Scores below clinical-significant levels (<29) of severe depression on the Beck Depression Inventory (BDI-II; Beck et al., 1996); 2) No current/history of mental health conditions; 3) No current/history of chronic pain; 4) None of the following conditions: cardiovascular disorder, Raynaud’s phenomenon, fainting, seizure, wounds on the hand/arm; 5) No frequent use of alcohol/tobacco; 6) Normal or corrected-to-normal vision; and 7) Right-handedness.

All participants provided their written informed consent and received a debriefing upon completion of the study. The study received ethical approval from the University of Hong Kong’s Human Research Ethics Committee for Non-Clinical Faculties (reference number: EA240016), and the protocols were pre-registered (NCT06866028, https://clinicaltrials.gov/study/NCT06866028).

#### 2.1.2 Randomized controlled design and blinding protocol

The investigation employed a randomized controlled trial design with participants allocated to either the active neurofeedback condition (experimental group) or a control condition (sham group), following recommended practices for neurofeedback studies (see supplementary table 1). The experimental group received real-time neurofeedback computed from their own brain activity during the session, while the sham group received feedback yoked from matched recordings of participants in the active condition; this ensured that participants in the sham group experienced similar visual feedback to the experimental group.

A partial double-blinding was implemented to minimize experimental bias and maintain methodological integrity. All participants were not aware of the sham control condition and completed the session under the belief that they were receiving training based on their real-time brain activity. The main experimenter was in direct contact with participants throughout the session and remained blinded to the group assignment to ensure the identical delivery of instructions to participants in both conditions. Another experimenter was responsible for monitoring the real-time neurofeedback interface and maintained knowledge of group assignments necessary for coordinating feedback delivery; as they had minimal interactions with the participants, the integrity of the blinding was preserved. At the end of the experiment, participants were debriefed about the inclusion of a sham control condition, and the group assignment was revealed to them upon request.

#### 2.1.3 Training procedures and paradigm

The experiment consisted of a single session, with assessments of pre-test and post-test outcomes conducted before and after the neurofeedback training. The session began with a pre-test period, where participants filled out demographic information and questionnaires.

Participants completed a training module, starting with an introduction to the neural basis of emotion regulation and the principles of neurofeedback. Then, participants were introduced to the concept of cognitive reappraisal and 3 sub-strategies: Positive Imagination (envisioning a more favorable outcome for current events), Alternative Explanation (generating alternative perspectives to reduce emotional intensity), and Reality Check (imagining the situation is fictional, such as a movie scene), in alignment with strategies implemented in other previous cognitive reappraisal-guided trainings (Zweerings et al., 2020; Keller et al., 2021). Participants completed a guided practice applying these sub-strategies towards exemplar negative images (depicting interpersonal violence, catastrophes, and threatening animals) to ensure familiarization with cognitive reappraisal.

Following this, participants received information about the subsequent neurofeedback training and standardized instructions to guide their reappraisals of the images (Jiang et al., 2025). Each trial began with a 2-4 second fixation, followed by a 1-second cue indicating participants to either REAPPRAISE or VIEW the upcoming negative image presented for 12 seconds. In VIEW trials, participants were instructed to look at the image attentively and experience their emotions naturally without attempting to control them; in REAPPRAISE trials, participants were asked to apply the reappraisal strategies learned from the training module to reduce their emotional reactivity towards the negative image. Following a 2-second blank interval after the image presentation, the screen displayed “How negative are you feeling right now?”, where participants made a rating from 1 (not at all negative) to 9 (very negative) within 4 seconds, and the trial ended with a 3-5 second jitter.

After setting up the fNIRS on participants, the formal training commenced, starting with a practice run (8 REAPPRAISE trials, 8 VIEW negative trials, and 8 VIEW neutral trials) without feedback, which also served as a functional localizer task. The main training consisted of 4 training runs (each run containing 8 VIEW negative and 8 REAPPRAISE trials), where feedback was administered for REAPPRAISE trials. The training concluded with a transfer run (8 REAPPRAISE trials and 8 VIEW negative trials) to evaluate regulation performance in the absence of feedback (see Fig. 1). All trials were pseudorandomized within each run to ensure the sham group received yoked feedback corresponding to REAPPRAISE trials during the training.

**Fig. 1.**
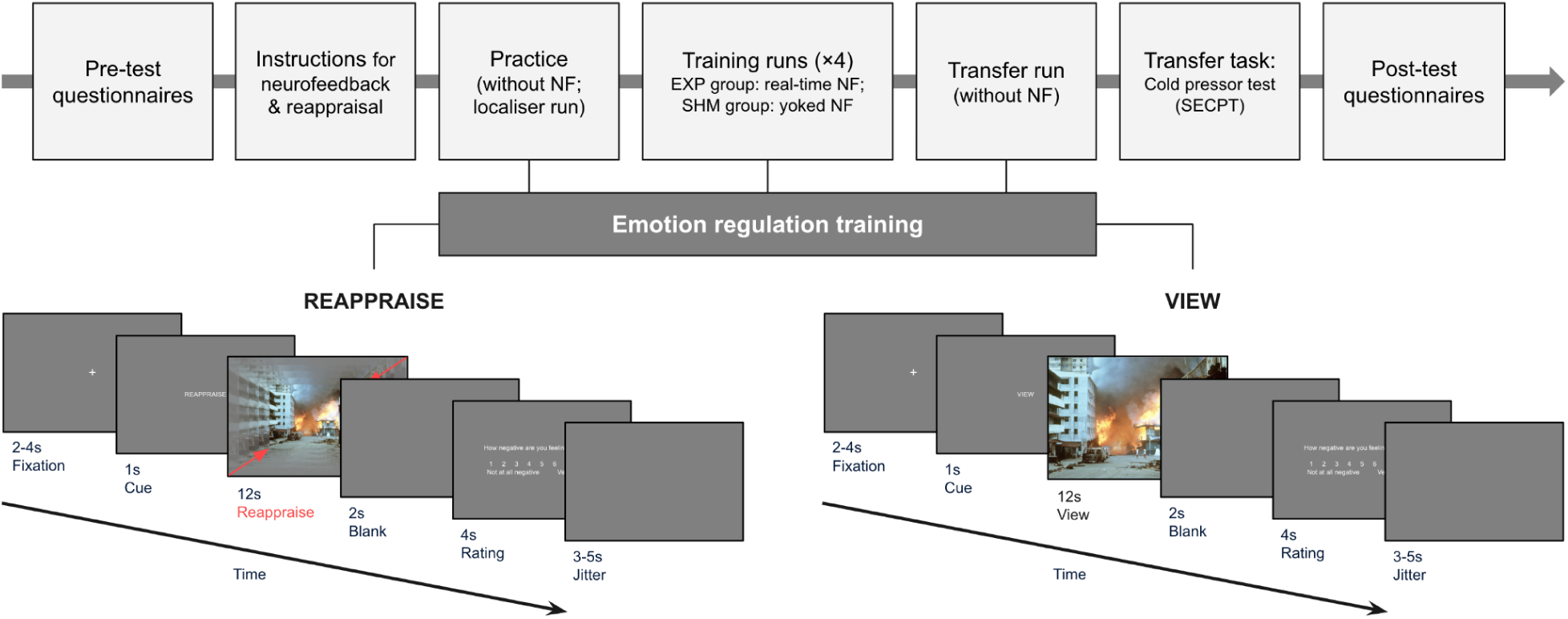
Experiment flow and task paradigm.

Participants then completed the SECPT (Schwabe et al., 2008), which involved the immersion of their non-dominant hand in cold water for a maximum of 3 minutes (i.e., 180 seconds), following standardized procedures (Schwabe & Schächinger, 2018). The task began with participants immersing their non-dominant hand in a warm water basin (35 ± 1.1°C) for 1 minute to normalize their hand temperature, followed by immersion in the cold water basin (1.7 ± 0.3°C) for the formal test. Participants were explicitly instructed to submerge their non-dominant hand in the cold water for as long as possible without knowledge of the 3-minute limit. To induce further stress through social evaluation, participants were also instructed to stare into the video monitor during the cold immersion, and were informed that their performance was being assessed by the experimenters and that the footage would be later analyzed. Participants rated on a visual analog scale from 0-100 their pain levels (0 = no pain; 100 = extremely severe pain), followed by stress levels (0 = no stress; 100 = extremely severe stress) before and after the test, which served as secondary outcome measures.

### 2.2 Materials

#### 2.2.1 Stimuli and task program

A series of databases (IAPS, Bradley & Lang, 2007; NAPS, Marchewka et al., 2014; SMID, Crone et al., 2018; and COMPASS, Weierich et al., 2019) were pooled, yielding 239 images (162 negative, 77 neutral) of animals, landscapes and people. These images were assessed by twenty independent raters for valence (1 = very negative, 9 = very positive) and arousal (1 = no response at all, 9 = very strong response) with the Self-Assessment Manikin (Bradley & Lang, 1994). 144 negative images were selected for the training, evenly distributed at random to VIEW (valence: *M* = 3.17, *SD* = 0.55) and REAPPRAISE (valence: *M* = 3.05, *SD* = 0.49) conditions; the valence of images did not significantly differ between both conditions (*p* = .158). 40 neutral images were selected (valence: *M* = 5.41, *SD* = 0.33; arousal: *M* = 3.64, *SD* = 0.33), 8 were presented in the practice run alongside negative images, and the remaining were presented during breaks between the training runs.

The experiment program was set up using PsychoPy v.2024.1.4 (Peirce et al., 2019). Triggers were sent from PsychoPy to the fNIRS data acquisition software via lab streaming layer, and from the acquisition software to the neurofeedback platform via TCP/IP networks; and feedback values from the neurofeedback platform were sent back to PsychoPy through interfacing with expyriment (Krause & Lindemann, 2014). For practice and transfer runs, images for VIEW and REAPPRAISE trials were presented at full height of the screen, and the width of the image was scaled to maintain its aspect ratio; this was also the default for VIEW trials during training runs.

#### 2.2.2 fNIRS data acquisition

fNIRS data was acquired with a NIRSport2 device and Aurora software from NIRx (Berlin, Germany). The NIRSport2 device recorded oxygenated hemoglobin (HbO) and deoxygenated hemoglobin (HbR) absorption at two wavelengths of light (760 and 850 nm). 16 light sources and 14 light detectors were used to create an interface of 50 long-separation channels covering prefrontal cortical regions (see Fig. 2). The placements for these channels were determined based on the existing templates and the fNIRS optodes’ location decider toolbox (Zimeo Morais et al., 2018), mapped onto standard 10–20 locations and with source-detector distances at around 3-cm. 8 short-separation channels were placed at lateral source locations. D01 was placed above the Nasion at 10% of the Nz–Iz distance in correspondence with FPz. Raw voltage data was acquired at a sampling rate of 5.09 Hz using the NIRx Aurora software.

**Fig. 2.**
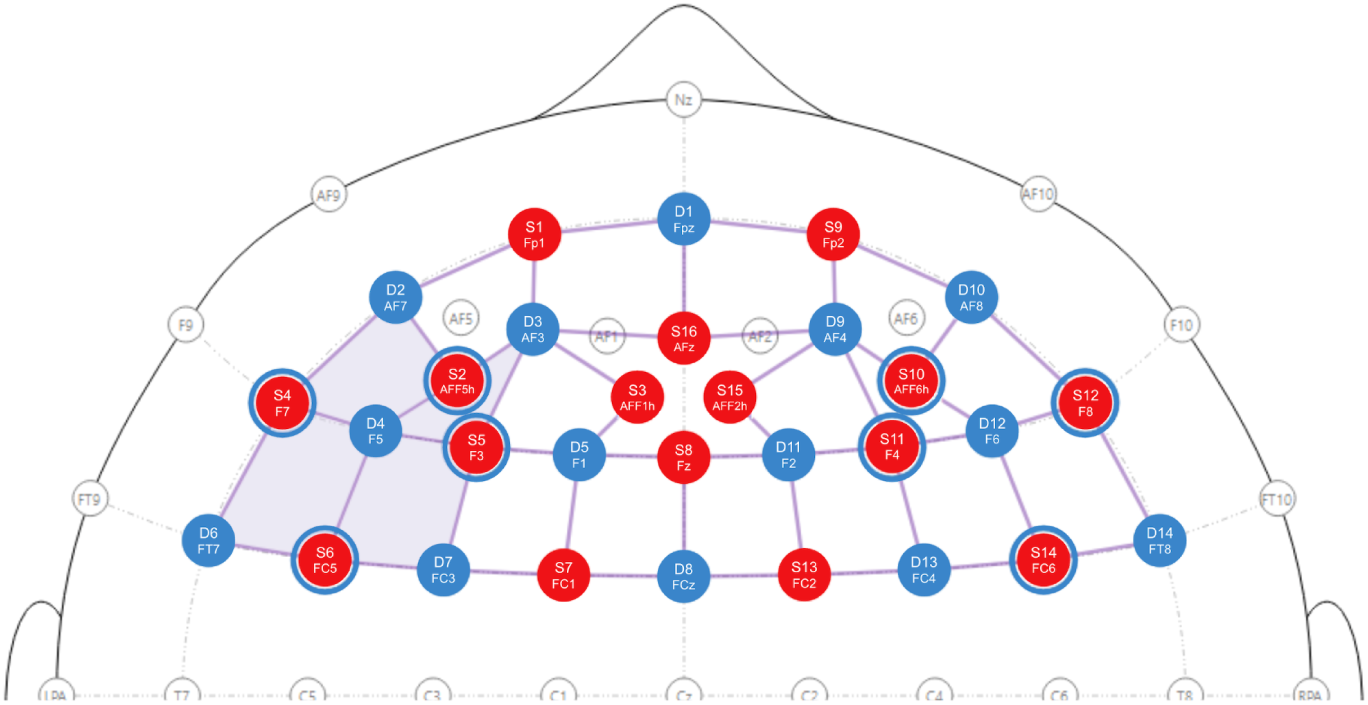
fNIRS cap configuration designed on 10–20 system with the ROI mask shaded

An *a priori* region-of-interest (ROI) channel mask was set at the left lateral PFC, consistent with previous literature suggesting reappraisal processes demonstrate stronger activations in left than right lateral PFC (Picó-Pérez et al., 2017; Silvers et al., 2015). These channels corresponded to automated anatomical labeling (AAL) regions (see supplementary table 2) covering the ventrolateral PFC (frontal inferior operculum, frontal inferior triangularis, frontal inferior orbitalis) and dorsolateral PFC (middle frontal gyrus, parts of superior frontal gyrus). This ROI mask guided the selection of individualized target channels during the functional localizer run for each participant, and further statistical comparisons to determine regional-specific effects.

#### 2.2.3 Real-time neurofeedback platform

Neurofeedback was administered using Turbo-Satori (Brain Innovation; Maastricht, Netherlands; Lührs & Goebel, 2017), a real-time analysis software for fNIRS. The software applies preprocessing parameters optimized for efficient computation to convert raw light intensity data to changes in HbO and HbR activity in real-time. The software interface displays block averages for conditions, enabling visual inspection of the signal quality and continuous monitoring of hemoglobin levels throughout the recording. During the practice and transfer runs, both groups had their real-time brain activity processed through the software. For the training runs, the experimental group received real-time feedback generated from their brain activity, while the sham control group received yoked feedback from a previous recording of a participant in the experimental group completing the same run.

The real-time processing protocol began with a 20-second baseline period to establish reference values for signal processing. An additional 10-second buffer period followed before task initiation to ensure consistent recording durations across all participants. Preprocessing steps were applied after raw wavelength conversion to hemoglobin concentrations in the following sequence: Linear detrending to remove offsets and slow drifts, then a moving average filter with high-pass (0.03 Hz) and low-pass (0.4 Hz) cutoffs to attenuate frequencies outside of the range of brain activity, and short separation correction (Goodwin et al., 2014) to regress out extracerebral noise captured by the short channels. The software computed *t*-tests for each channel within 20-second windows across the trials, and the scale factor was set to 500 to ensure adequate sensitivity for detecting activity changes while maintaining consistent feedback scaling across all sessions.

### 2.3 Neurofeedback Training

#### 2.3.1 Practice run and functional localizer

The practice run served as a functional localizer task to select channels appropriate for neurofeedback, with the contrast set to REAPPRAISE > VIEW (negative). The real-time processing platform continuously computed *t*-test comparisons for each channel and displayed average waveform plots for both conditions throughout the recording. Individualized target channels were determined by identifying those that consistently produced high *t*-test values (at an uncorrected threshold of *p* < .05) and showed increased HbO activity during REAPPRAISE trials, indicative of brain activity coupled to the regulation mechanism of interest.

#### 2.3.2 Training runs

Continuous feedback was relayed during REAPPRAISE trials by calculating values based on averaged HbO activity at target channels, subtracting the preceding baseline measurements. These values, which typically range from -1 to 1 on Turbo-Satori, were multiplied by 10 to convert them into appropriate scaling values for the feedback. For the first 2 seconds of the presentation duration, the images were scaled at the default size. After that, feedback was updated continually every second with a looming size paradigm, based on a scaling factor 𝑆 calculated using 𝑆 = 1/((𝑣/4) + 1), where 𝑣 represents the feedback value (1≤𝑣≤10). The image height was directly scaled by 𝑆 and its width was proportionally adjusted to maintain the original aspect ratio; the image shrunk more visibly at lower activation levels to encourage initial regulation, and gained more stability in size as activation levels increased to signal achieved control.

#### 2.3.3 Transfer run

The emotion regulation training ended with a transfer run where, just like in the practice run, participants did not receive feedback for REAPPRAISE trials. This served to determine whether neural regulation was maintained following the training runs in the absence of feedback, providing an assessment of whether participants had internalized the regulatory strategies and could sustain brain activity changes independently.

### 2.4 Analyses

#### 2.4.1 Offline fNIRS preprocessing and analysis

The fNIRS data underwent preprocessing and analysis on MATLAB using the NIRS Brain AnalyzIR toolbox (Santosa et al., 2018). Raw light intensity measurements were first trimmed to retain the 30-second baseline period preceding each recording, then downsampled from the original acquisition rate to 4 Hz to complement the subsequent prewhitening algorithm in the first-level analysis. The intensity data was converted to optical density values, followed by low-pass filtering at 0.5 Hz to eliminate high-frequency noise artefacts. Optical density measurements were then transformed into HbO and HbR concentration changes using the modified Beer-Lambert law with partial pathlength factor set to 0.1.

The general linear model (GLM) analyses employed the autoregressive iteratively reweighted least-squares method (AR-IRLS; Barker et al., 2013). For each channel, parameters were initialized with ordinary least squares estimates; then, an optimal-order autoregressive model was applied to prewhiten the data, addressing serial correlations present from systemic physiology (i.e., cardiac, respiratory and blood pressure oscillations). The model was then solved using iteratively reweighted least squares to minimize the influence of outliers from motion artefacts, effectively controlling for type I errors. In addition, short-separation regression with orthonormal decomposition (Santosa et al., 2020) was implemented to further correct for noise originating from extracerebral components in convolving the hemodynamic response. The resulting parameter estimates were extracted for second-level analyses.

One participant’s fNIRS data from the practice run was due to technical failure. As a data quality control step, scalp-coupling indices (SCIs; Pollonini et al., 2016) were computed on raw signals of long-separation channels for all recordings. For each participant, channels with SCIs below 0.8 across more than half of the session were excluded in subsequent statistical analyses of channel activity.

#### 2.4.2 Statistical analyses

All statistical analyses were performed using RStudio (v.2025.5.0.496).

Negativity ratings were screened for quality, excluding participants with excessive missing ratings (>50%) or insufficient response variability (*SD* < 1). Based on these criteria, four participants were removed: one had missing data as a result of technical errors (p03) and three were excluded due to invariant responses (p04, p08, p32), resulting in the exclusion of two participants from each group. Another participant (p57) missed ratings during the first half of the practice run, resulting in the exclusion of REAPPRAISE trials due to a low number of ratings.

### 2.5 Outcomes of the Training

#### 2.5.1 Primary outcomes: Activations in target channels

The present study focused on the specific effects of neurofeedback training, therefore, the primary outcomes involved comparing activations at individualized target channels. Specifically, we analyzed parameter estimates for REAPPRAISE trials averaged across the target channels using mixed ANOVAs comparing the experimental vs. sham groups across training runs (run 1 vs. 2 vs. 3 vs. 4) and pre- and post-training (practice run vs. transfer run). Significant effects were followed by Bonferroni-Holm corrected post hoc comparisons.

In addition, we computed channel-wise comparisons across the training to assess if an effect of neurofeedback training could be pinpointed to a specific region of the lateral PFC, reporting channels with upwards activations (i.e., where parameter estimates positively increased beyond 0 throughout the period) that illustrate significant group or interaction effects.

#### 2.5.2 Secondary outcomes: Socially-evaluated cold pressor test

Stress and pain ratings collected before and after completion of the SECPT were analyzed using mixed ANOVAs comparing ratings made by both groups before and after completion of the procedure to verify the success of the induction procedure (i.e., significant time effects). To assess our secondary outcome, we computed one-tailed *t*-tests comparing post-test ratings of stress and pain levels between the groups, based on an *a priori* directional hypothesis that the training would result in lower ratings in the neurofeedback group than the sham group.

#### 2.5.3 Behavioral outcomes and control measures

To determine if neurofeedback facilitated enhanced regulation that coincided with behavioural outcomes, we turned to self-report measures of anxiety and affect (valence and arousal) collected at pre- and post-training timepoints. Participants made ratings of anxiety (on a VAS from 0-100; 0 = not at all anxious; 100 = extremely anxious) and affect (valence and arousal; rated using the Affect Grid by Russell et al., 1989) at pre-training and post-training timepoints. We also compared negativity ratings made for REAPPRAISE and VIEW trials across the training runs, and between the practice and transfer runs. For each measure, we conducted mixed ANOVAs with group as the between-subjects factor and timepoint as the within-subjects factor, followed by planned comparisons to assess group differences at each timepoint.

Given the between-groups design, we assessed different measures to control for group differences: Perceived Stress Scale (PSS; Cohen et al., 1983); Kessler Psychological Distress Scale (K10; Kessler et al., 2002); State–Trait Anxiety Inventory (STAI; Spielberger et al., 1983); Pain Sensitivity Questionnaire (PSQ; Ruscheweyh et al., 2009); Neuroticism subscale of the NEO-FFI (Costa & McCrae, 1992); Self-Efficacy Scale (SES; Sherer et al., 1982).

Lastly, we assessed blinding efficacy through multiple measures, including participants’ ratings of their perceived ability to control brain activity and success in applying cognitive reappraisal strategies at pre- and post-training timepoints. Participants were asked to rank the sub-strategies they applied from highest to lowest after the training ended. At the end of the experiment, participants were asked whether they believed they received real neurofeedback, and rated their confidence levels on their judgment.

## 3 Results

In line with our pre-registration, 60 participants (*M* = 20.13, *SD* = 2.33) were recruited. The experimental and sham groups did not significantly differ in psychosocial factors (see Table 1).

**Table 1.** Demographics information for the groups.

| | **Experimental group**<br>$N = 30$ (11 males) | **Sham group**<br>$N = 30$ (13 males) | ***t*-test** |
| --- | --- | --- | --- |
| **Age** | $20.07 \pm 2.26$ | $20.20 \pm 2.44$ | $p = .827$ |
| **BDI-II** | $11.03 \pm 6.84$ | $10.80 \pm 7.19$ | $p = .898$ |
| **PSS** | $18.23 \pm 4.78$ | $18.60 \pm 4.72$ | $p = .766$ |
| **K10** | $21.17 \pm 4.94$ | $21.13 \pm 6.17$ | $p = .982$ |
| **STAI-State** | $39.40 \pm 8.31$ | $39.10 \pm 7.63$ | $p = .885$ |
| **STAI-Trait** | $45.43 \pm 6.53$ | $45.03 \pm 7.11$ | $p = .821$ |
| **PSQ** | $65.93 \pm 16.83$ | $65.30 \pm 21.52$ | $p = .899$ |
| **Neuroticism (NEO-FFI)** | $36.27 \pm 6.58$ | $36.20 \pm 5.77$ | $p = .967$ |
| **SES** | $76.93 \pm 11.01$ | $72.90 \pm 10.28$ | $p = .148$ |

### 3.1 Primary outcome: Training success on the neural level

#### 3.1.1 Neurofeedback effect across the training runs

Target channel activations differed significantly between groups across the training, *F*(1, 58) = 9.20, *p* = .004, *ηp²* = .14 (see Fig. 3), with the experimental group showing higher activations than the sham group for run 1, *t*(58) = 2.19, *p* = .033, Cohen’s *d* = .57 (experimental: *M* = 2.46, *SE* = 0.72; sham: *M* = 0.12, *SE* = 0.80), run 2, *t*(58) = 2.62, *p* = .011, Cohen’s *d* = .69 (experimental: *M* = 2.24, *SE* = 0.79; sham: *M* = -0.41, *SE* = 0.64), and run 3, *t*(58) = 2.64, *p* = .011, Cohen’s *d* = .69 (experimental: *M* = 3.19, *SE* = 1.25; sham: *M* = -0.62, *SE* = 0.74), and marginally for run 4, *t*(58) = 2.00, *p* = .0503, Cohen’s d = .53 (experimental: *M* = 4.91, *SE* = 0.90; sham: *M* = -0.61, *SE* = 1.03). No main effect of run (*F*(3, 174) = 0.35, *p* = .793), or interaction effect (*F*(3, 174) = 0.49, *p* = .688) was present.

**Fig. 3.**
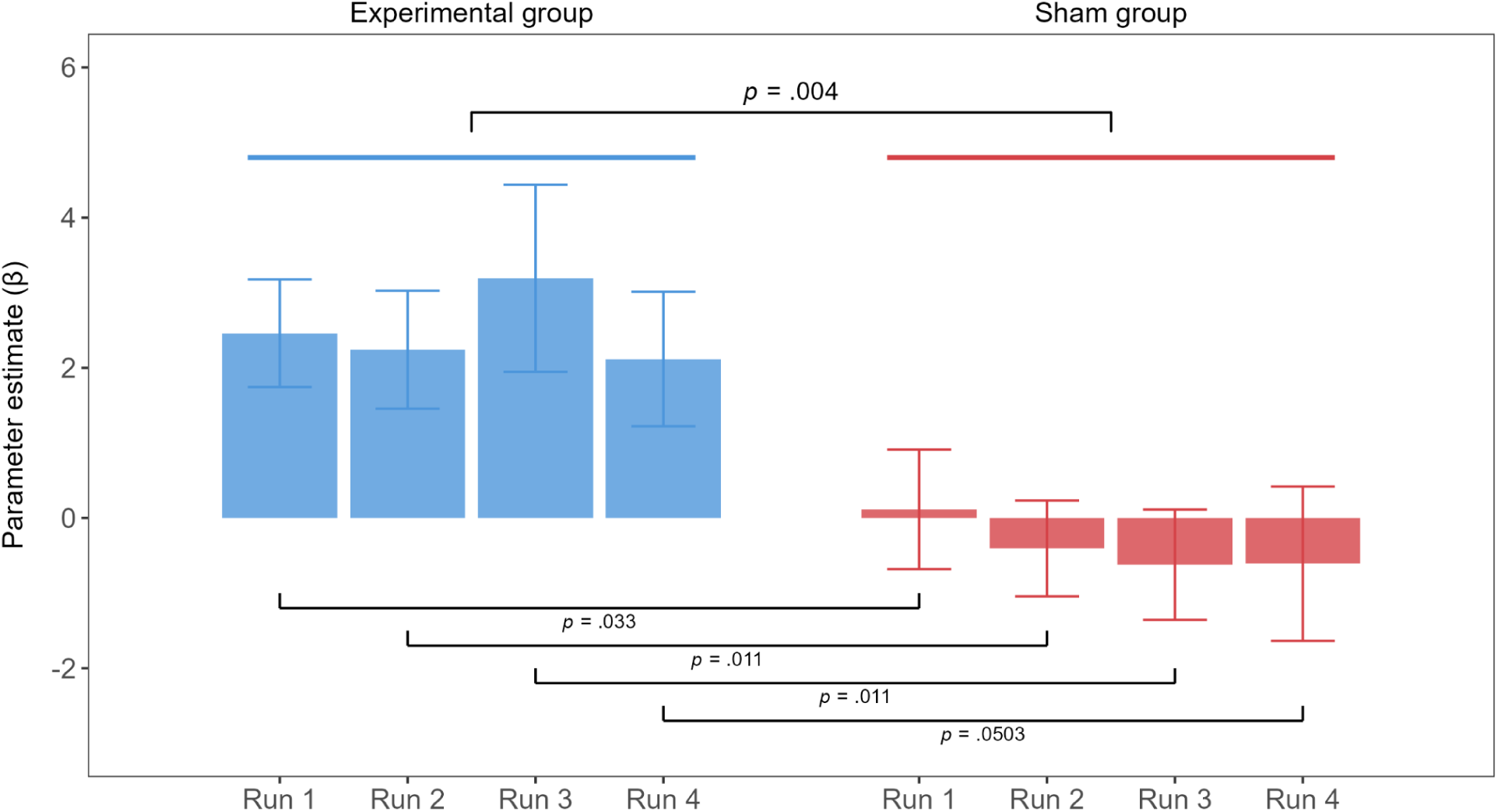
Parameter estimates of brain activation at individualized target channels across the training

#### 3.1.2 Comparison between practice and transfer runs

Comparisons of target channel activations in the practice and transfer runs revealed a significant effect of group, *F*(1, 57) = 6.46, *p* = .014, *ηp²* = .10. While the experimental and sham groups did not differ in the practice run, *t*(57) = 1.33, *p* = .190 (experimental: *M* = 2.79, *SE* = 0.70; sham: *M* = 1.28, *SE* = 0.90), the experimental group had higher activations than the sham group for the transfer run, *t*(57) = 2.64, *p* = .011, Cohen’s *d* = .70 (experimental: *M* = 2.40, *SE* = 0.90; sham: *M* = -0.39, *SE* = 0.55). No main effect of run (*F*(1, 57) = 2.16, *p* = .147) or interaction effect (*F*(1, 57) = 0.83, *p* = .367) was present.

#### 3.1.3 Channel specificity of neurofeedback training

During the training, there was a significant interaction between group and run for channel S4-D6 within the ROI mask, corresponding to the left inferior frontal gyrus, *F*(2.19, 122.87) = 4.51, *p* = .048, *ηp²* = .05, with a marginally significant effect of run, *F*(2.19, 122.87) = 2.63, *p* = .071, *ηp²* = .05, but no effect of group, *F*(1, 56) = 2.50, *p* = .120. Post hoc comparisons revealed a significant difference during run 3, *t*(56) = 2.83, *p* = .006, Cohen’s *d* = .76, between the experimental group (*M* = 6.54, *SE* = 1.43) and sham group (*M* = 0.20, *SE* = 1.68) (see Fig. 4).

**Fig. 4.**
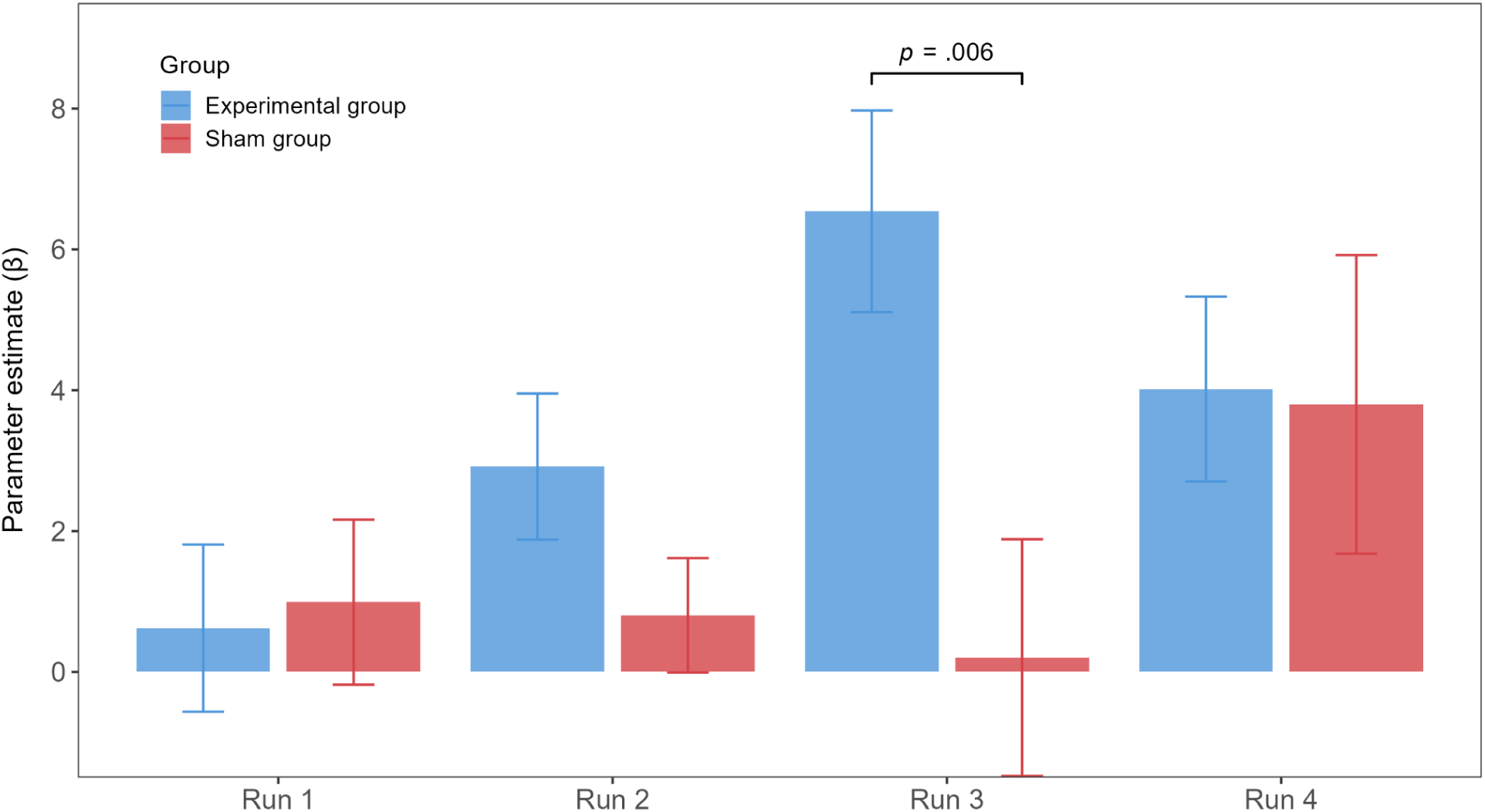
Parameter estimates of brain activation at channel S4-D6 (inferior frontal gyrus) across the training

Comparing the practice and transfer runs, there was a significant group effect present, *F*(1, 55) = 5.47, *p* = .023, *ηp²* = .09, with no significant effect of run, *F*(1, 55) = 0.63, *p* = .431, or interaction effect, *F*(1, 55) = 0.07, *p* = .788. A marginally significant difference was present at the transfer run, *t*(55) = 1.93, *p* = .059, Cohen’s *d* = .52, between the experimental group (*M* = 3.31, *SE* = 0.97) and sham group (*M* = 0.18, *SE* = 1.14).

### 3.2 Secondary outcomes from the stress induction procedure

#### 3.2.1 Stress regulation

A mixed ANOVA comparing stress ratings before and after the SECPT revealed a significant main effect of timepoint, *F*(1, 58) = 65.65, *p* < .001, *ηp²* = .53, confirming that the procedure successfully induced stress across participants, with higher stress reported post-induction (*M* = 56.12, *SE* = 3.64) than pre-induction (*M* = 23.33, *SE* = 2.65). The group effect, *F*(1, 58) = 1.95, *p* = .168, and interaction effect, *F*(1, 58) = 1.62, *p* = .208, were not significant.

At baseline, stress levels were comparable between the experimental group (*M* = 22.70, *SE* = 4.13) and sham group (*M* = 23.97, *SE* = 3.39), *t*(58) = -0.24, *p* = .407 (one-tailed). However, following completion of the procedure, stress levels were significantly lower for the experimental group (*M* = 50.33, *SE* = 5.14) than the sham group (*M* = 61.90, *SE* = 4.49), *t*(58) = -1.70, *p* = .048, Cohen’s *d* = -.45 (one-tailed; see Fig. 5a). Comparing the magnitude of stress levels between conditions indicated that there was a 27.2% reduction in stress in the experimental group compared to the sham condition.

**Fig. 5.**
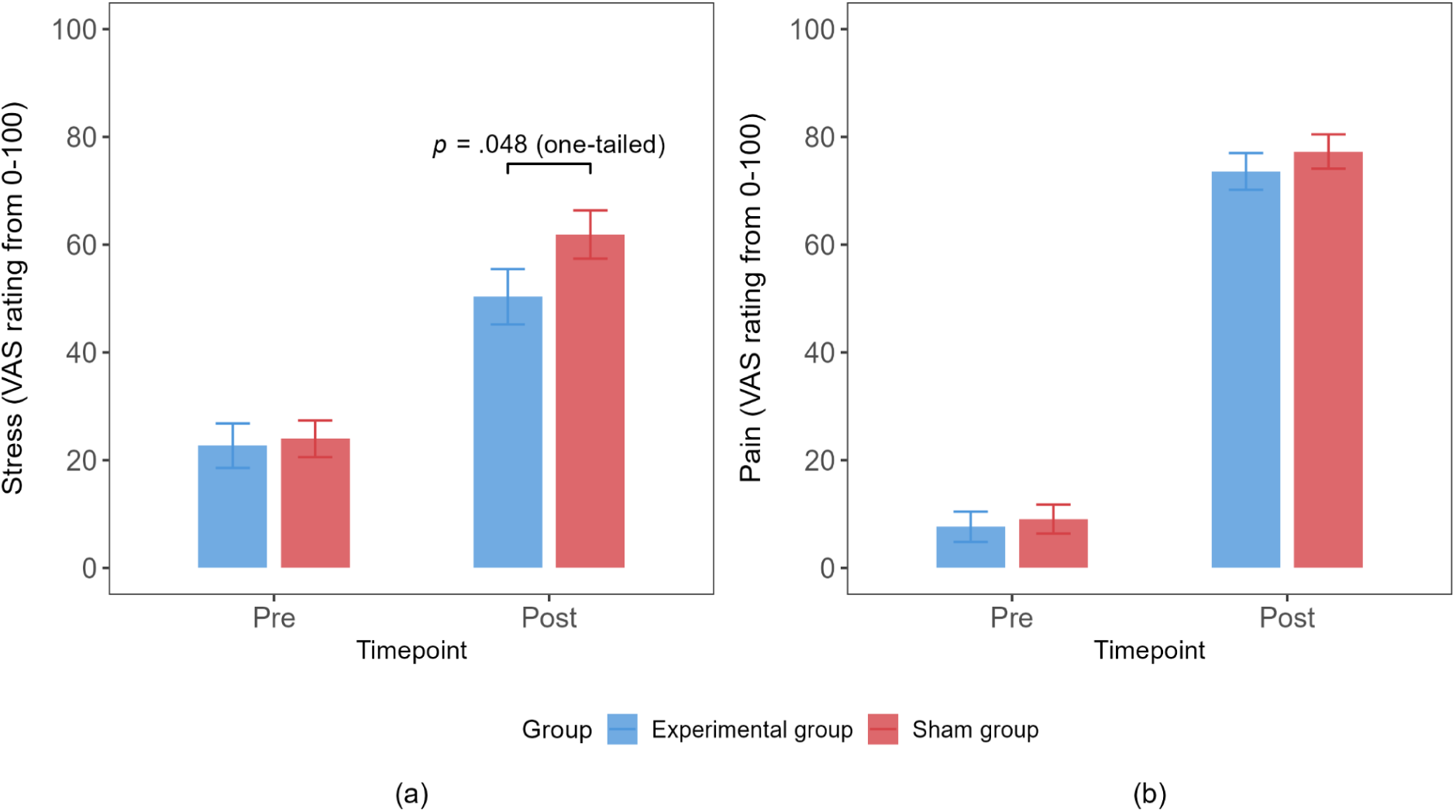
Secondary outcomes: (a) stress regulation and (b) pain regulation from the SECPT

#### 3.2.2 Pain regulation

For pain ratings, the mixed ANOVA revealed a significant main effect of timepoint, *F*(1, 58) = 664.36, *p* < .001, *ηp²* = .92, with substantially higher pain reported post-induction (*M* = 75.45, *SE* = 2.33) than pre-induction (*M* = 8.35, *SE* = 1.93). Neither the main effect of group, *F*(1, 58) = 0.56, *p* = .456, nor interaction effect, *F*(1, 58) = 0.19, *p* = .665, reached significance. Self-reported pain levels did not differ significantly between the groups (see Fig. 5b) at baseline, *t*(58) = -0.369, *p* = .357 (one-tailed; experimental: *M* = 7.63, *SE* = 2.81; sham: *M* = 9.07, *SE* = 2.69), nor after the SECPT, *t*(58) = -0.79, *p* = .216 (one-tailed; experimental: *M* = 73.60, *SE* = 3.42; sham: *M* = 77.30, *SE* = 3.19). Additionally, participants submerged their hands in cold water for similar durations, *t*(57) = 0.824, *p* = .413 (two-tailed; experimental: *M* = 153.3 s, *SE* = 10.6 s; sham: *M* = 140.3 s, *SE* = 11.6 s).

### 3.3 Behavioral outcomes from training

#### 3.3.1 Pre- and post-training ratings

Analysis of anxiety levels with a mixed ANOVA revealed a marginally significant interaction effect between timepoint and group, *F*(1, 58) = 3.02, *p* = .088, *ηp²* = .05. No main effect of group (*F*(1, 58) = 0.26, *p* = .612) or timepoint (*F*(1, 58) = 0.04, *p* = .844) was present. Comparisons confirmed that the two groups did not differ in anxiety levels pre-training, *t*(58) = 0.319, *p* = .376 (one-tailed; experimental: *M* = 29.80, *SE* = 4.19; sham: *M* = 28.00, *SE* = 3.79). Notably, anxiety levels diverged post-training, *t*(58) = -1.309, *p* = .098, Cohen’s *d* = .34 (one-tailed), with anxiety levels diminishing in the experimental group (*M* = 26.03, *SE* = 3.62) while anxiety levels elevated in the sham group (*M* = 32.73, *SE* = 3.62) (see Fig. 6a).

**Fig. 6.**
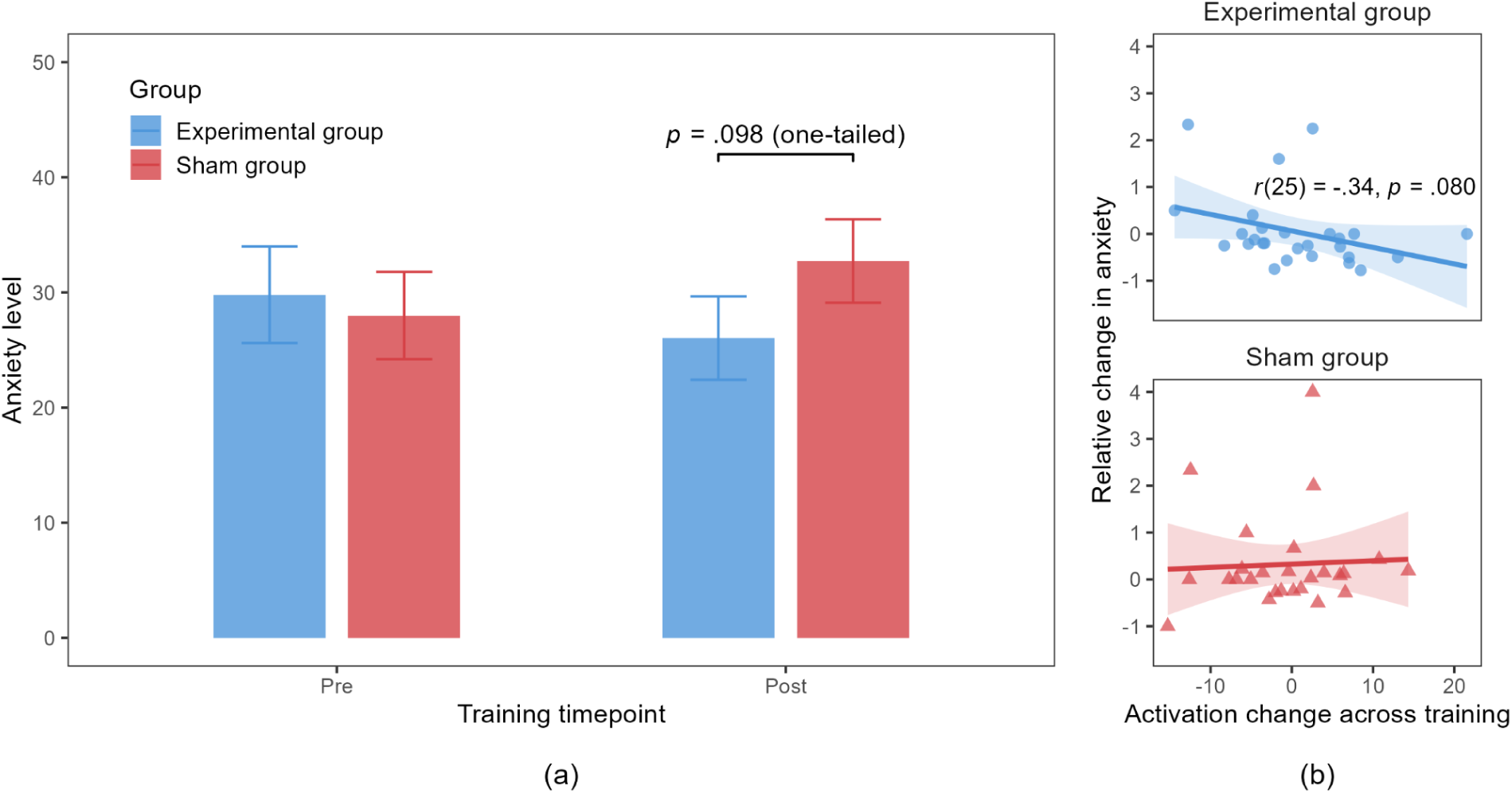
(a) Change in anxiety before and after training, (b) correlated with changes in brain activation across training

As these changes occurred within the training, we sought to examine whether this change in anxiety was associated with changes in brain activity observed at target channels. We calculated relative change in anxiety (post-training minus pre-training anxiety, divided by pre-training anxiety) and quantified neural activation change as the difference in parameter estimates between the later training phase (runs 3 & 4) and the initial training phase (runs 1 & 2). After excluding participants (experimental: *N* = 3; sham: *N* = 4) with non-computable data points (due pre-training anxiety ratings of 0) and statistical outliers falling outside ± 3 *SD*, analysis revealed a marginally significant negative correlation between activation change and anxiety reduction in the experimental group (see Fig. 6b), Pearson’s *r*(25) = -.34, *p* = .080; no significant association was observed in the sham group, Pearson’s *r*(24) = .05, *p* = .812.

Valence ratings analysed with a mixed ANOVA showed a marginally significant effect of timepoint, *F*(1, 58) = 3.37, *p* = .071, *ηp²* = .05, but no effect of group, *F*(1, 58) = 0.36, *p* = .550 and no interaction effect, *F*(1, 58) = 1.13, *p* = .292. Between-group comparisons showed no significant differences in valence ratings pre-training, *t*(58) = -1.136, *p* = .260 (experimental: *M* = 5.90, *SE* = 0.27; sham: *M* = 6.40, *SE* = 0.35), nor post-training, *t*(58) = .11, *p* = .916 (experimental: *M* = 5.70, *SE* = 0.31; sham: *M* = 5.65, *SE* = 0.35).

Arousal ratings showed no significant effects for group, *F*(1, 58) = 0.08, *p* = .776, timepoint, *F*(1, 58) = 0.05, *p* = .828, or interaction effect, *F*(1, 58) = 0.82, *p* = .369. Consistent with these ANOVA results, no significant differences were observed between groups at pre-training, *t*(58) = .23, *p* = .822 (experimental: *M* = 4.40, *SE* = 0.29; sham: *M* = 4.28, *SE* = 0.42), or post-training, *t*(58) = -.72, *p* = .473 (experimental: *M* = 4.10, *SE* = 0.34; sham: *M* = 4.47, *SE* = 0.37).

#### 3.3.2 Negativity ratings

For REAPPRAISE trials across the training runs, we observed a significant effect of time, *F*(3, 162) = 7.90, *p* < .001, *ηp²* = .13. A significant decrease in negativity ratings was observed in run 4 (*M* = 3.68, *SE* = 0.19) compared to run 1 (*M* = 4.18, *SE* = 0.18), *t*(54) = -3.68, *p* = .003, Cohen’s *d* = -1.00, run 2 (*M* = 4.01, *SE* = 0.19), *t*(54) = -2.88, *p* = .023, Cohen’s *d* = -.79, and run 3 (*M* = 4.20, *SE* = 0.21), *t*(54) = -4.57, *p* < .001, Cohen’s *d* = -1.2. There was no group effect, *F*(1, 54) = 0.14, *p* = .706, or interaction effect, *F*(3, 162) = 0.48, *p* = .695.

Comparing the practice and transfer runs, there was similarly a significant effect of time, *F*(1, 53) = 8.50, *p* = .005, *ηp²* = .14 (practice: *M* = 4.28, *SE* = 0.19; transfer: *M* = 3.79, *SE* = 0.20), with no group effect, *F*(1, 53) = 0.02, *p* = .896, or interaction effect, *F*(1, 53) = 0.27, *p* = .605. Further analyses indicated ratings decreased between practice and transfer runs for the experimental group, *t*(53) = 2.45, *p* = .018, Cohen’s *d* = .67 (practice: *M* = 4.29, *SE* = 0.27; transfer: *M* = 3.75, *SE* = 0.29), and not the sham group, *t*(53) = 1.68, *p* = .099 (practice: *M* = 4.26, *SE* = 0.26; transfer: *M* = 3.84, *SE* = 0.29).

For VIEW trials across the training runs, there was a significant effect of time, *F*(3, 162) = 13.23, *p* < .001, *ηp²* = .20. Run 3 (*M* = 4.79, *SE* = 0.22) showed significantly lower ratings compared to run 1 (*t*(54) = 5.08, *p* < .001, Cohen’s *d* = 1.38; run 1: *M* = 5.38, *SE* = 0.22), run 2 (*t*(54) = 5.83, *p* < .001, Cohen’s *d* = 1.59; run 2: *M* = 5.43, *SE* = 0.23), and run 4 (*t*(54) = 5.12, *p* < .001, Cohen’s *d* = 1.39; run 4: *M* = 5.30, *SE* = 0.23). No group effect, *F*(1, 54) = 2.78, *p* = .101, or interaction effect, *F*(3, 162) = 1.41, p = .243, were present.

There was a marginally significant interaction effect for practice versus transfer runs, *F*(1, 54) = 2.92, *p* = .093, *ηp²* = .05; however, differences were not significant for the practice, *t*(54) = -0.54, *p* = .593 (experimental: *M* = 4.94, *SE* = 0.26; sham: *M* = 5.15, *SE* = 0.31), nor transfer run, *t*(54) = -1.65, *p* = .105 (experimental: *M* = 4.76, *SE* = 0.26; sham: *M* = 5.47, *SE* = 0.34). There was no effect of time, *F*(1, 54) = 0.22, *p* = .641, or effect of group, *F*(1, 54) = 1.40, *p* = .242.

### 3.4 Control measures: Blinding efficacy and training outcomes

A chi-square analysis revealed no significant association between group assignment and participants’ beliefs of whether they believed they received real neurofeedback or not, *χ²*(1, *N* = 60) = 0.577, *p* = .448, *φ* = .10. In the experimental group, 90% participants (*N* = 27) believed they received real neurofeedback compared to 83.3% (*N* = 25) in the sham group. Confidence ratings regarding these beliefs did not differ between the experimental (*M* = 6.23, *SD* = 1.72) and sham (*M* = 6.17, *SD* = 1.95) groups, *t*(58) = 0.14, *p* = .889.

We assessed training outcomes to determine if blinding procedures successfully prevented confound effects. There were no differences in post-training ratings of perceived control over brain activity, *t*(58) = -1.51, *p* = .137 (experimental: *M* = 5.90, *SD* = 1.30; sham: *M* = 6.40, *SD* = 1.28), though pre-training expected control ratings were higher for the sham group (*M* = 6.80, *SD* = 0.89) than the experimental group (*M* = 6.13, *SD* = 1.14), *t*(58) = -2.53, *p* = .014. Similarly, in terms of expected success in applying strategies, the sham group had higher ratings (*M* = 6.57, *SD* = 1.04) than the experimental group (*M* = 6.07, *SD* = 1.01), *t*(58) = -1.88, *p* = .064; though post-training, both groups had similar ratings of perceived success, *t*(58) = -1.13, *p* = .262 (experimental: *M* = 6.53, *SD* = 0.68; sham: *M* = 6.80, *SD* = 1.10). This confirms the observed neurofeedback effects were not due to expectation effects nor better understanding of strategies.

As exploratory analyses, we assessed participants’ use of cognitive reappraisal by comparing rankings of the three sub-strategies with Mann-Whitney U tests. For Positive Imagination, the experimental group assigned higher ranks (*Mdn* = 1) than the sham group (*Mdn* = 2), *U* = 300, *p* = .013, *r* = .33. For Alternative Explanation, the sham group assigned higher ranks (*Mdn* = 1) than the experimental group (*Mdn* = 2), *U* = 345, *p* = .085, *r* = -.23. Rankings for Reality Check did not differ between the groups (experimental: *Mdn* = 3; sham: *Mdn* = 3), *U* = 376, *p* = .106, *r* = -.16.

## 4 Discussion

The present study investigated the efficacy of enhancing stress regulation with fNIRS-based neurofeedback through a methodologically rigorous pre-registered double-blind, randomized sham-controlled design. Our main objectives were to determine whether real-time neurofeedback with explicit regulation using cognitive reappraisal strategies would increase activity at individualized lateral prefrontal target channels compared to a sham control condition (primary outcome) and to determine whether neurofeedback training effects translate into enhanced and domain-specific regulation of stress in the absence of feedback and under ecologically valid stressors (secondary outcome). This addressed critical prerequisites essential for translating laboratory-based neurofeedback into real-world applications: the maintenance of regulatory effects in the absence of feedback, and transfer of acquired regulation skills to novel stressful contexts. Our findings present converging evidence of training-induced increased activity in lateral prefrontal regions and successful transfer into improved stress regulation in a subsequent ecologically valid challenge. While the effects of the training were flanked by a trend of decreased anxiety levels after training, the groups did not differ in the pain experience during the stress task or emotion regulation success during the training, suggesting maintenance and stress-specific effects of the training.

### 4.1 Neurofeedback training effect

Our analysis revealed evidence that fNIRS-based neurofeedback successfully increased activations in targeted prefrontal regions. A significant group effect arose from the training: the experimental group showed consistently higher activations at targeted channels compared to the sham group. Notably, this activation difference emerged rapidly, with participants in the experimental group demonstrating efficient learning as early as the first run, mirroring findings from a recent fMRI neurofeedback study employing interoceptive strategies to enhance insula regulation (Zhang et al., 2023). This enhancement persisted through to the transfer run where participants no longer received feedback, demonstrating a genuine training effect. The differential pattern of activity is particularly noteworthy as it indicates that the increased activity observed in the experimental group was not merely a transient effect: real-time neurofeedback provided unique information that allowed participants to more effectively engage target regulatory regions, an effect not attributable to general engagement, reappraisal training or other confounding factors.

In comparing channel-wise activations across the training, our analyses identified channel S4-D6, corresponding to the inferior frontal gyrus within the lateral PFC, as showing pronounced upregulation preferentially in the experimental group. As participants were able to optimize engagement of this key regulatory region, this suggests an efficient matching between the applied reappraisal strategies and the individualized lateral PFC target regions. The lateral PFC is well-established within emotion regulation networks (Kohn et al., 2014; Etkin et al., 2015), and our finding aligns with studies demonstrating its reliable activation during volitional regulation (Tupak et al., 2014; Morawetz et al., 2017). This specificity highlights a critical advantage of neurofeedback as an intervention approach: its ability to selectively target and boost activity in specific neural regions central to cognitive control processes fundamental to stress regulation.

The efficiency to which participants utilized feedback to upregulate activity at target channels, coupled with a channel-specific effect observed within the lateral PFC ROI, demonstrates the potential of this technique for strengthening regulatory neural circuits, offering a targeted dimension beyond conventional cognitive training approaches alone. Additionally, the absence of group differences in measures of perceived control over brain activity and success in applying strategies further confirms that these neurofeedback effects reflect genuine neural regulation rather than expectancy effects or differences in self-efficacy. Together, these findings provide strong evidence that fNIRS-based neurofeedback provides a precision-guided approach for individuals to implement regulatory strategies effectively.

### 4.2 Transferable effects of the neurofeedback training

To assess the effect of neurofeedback beyond the training session, participants completed a standardized cold pressor test, with stress and pain regulation serving as secondary outcomes. The cold water immersion effectively induced pain levels comparably between the two groups, confirming the task’s effectiveness as an ecologically valid and strong stressor. Critically, while participants in both groups began with nearly identical baseline stress levels and immersed their hands for comparable durations, the experimental group reported lower stress levels compared to the sham group following the cold pressor test. This pattern illustrates a domain-specific rather than general regulatory effect, suggesting that neurofeedback training specifically enhanced stress regulation capabilities without altering pain perception, consistent with a previous study that demonstrated improved empathic responding in the experimental group compared to the control group (Yao et al., 2016). These findings demonstrate that neurofeedback not only enhances prefrontal control engagement beyond standard emotion regulation training, but also promotes adaptive regulatory strategies that transfer to novel stressful contexts.

The enhanced stress regulation demonstrated by the experimental group was further reflected in anxiety levels measured throughout the training. While both groups reported similar anxiety levels at baseline, following training, the experimental group exhibited diminished anxiety, while anxiety levels in the sham group increased. Moreover, the relative change in anxiety post-training compared to pre-training significantly correlated with activation changes across the training in the experimental group, while no such relationship was observed in the sham group. This pattern is consistent with previous research training prefrontal–limbic connectivity in healthy subjects, who likewise observed reductions in anxiety from the training (Zhao et al., 2019). These findings suggest that neurofeedback training provided a protective effect against anxiety levels that may accumulate from the extensive training, potentially through enhanced neural regulation capabilities.

### 4.3 Effects of neurofeedback training on behavioral outcomes

Despite successfully enhancing prefrontal activation and improving regulation towards stress reactivity, the neurofeedback training did not produce differential effects on emotion processing during the training itself. Valence levels decreased between pre-training and post-training timepoints comparably between both groups, and no significant changes in arousal were observed in either group. For negativity ratings, a significant decrease was observed from the practice to transfer run for the experimental group and not the sham group for REAPPRAISE trials; however, no group differences were found within the training runs for REAPPRAISE and VIEW trials. This altogether suggests a dissociation between neural recruitment of regulatory regions and perceptible changes in subjective affect or negativity ratings within the training timeframe. In spite of the absence of concurrent behavioral improvements, regulatory adaptations from the training were functionally significant in the transfer task, where enhanced stress regulation within the neurofeedback group was observed.

Several possibilities may explain why the training affected regulation of stress following the training while producing minimal effects on subjective emotional responses during the training. Neurofeedback effects may manifest primarily after the training, though this remains largely unexplored since the temporal onset of effects from single-session neurofeedback interventions is particularly understudied. While multi-session protocols potentially yield stronger or more consistent effects (Thibault et al., 2018), the outcomes of single-session training remains systematically uncharacterised, with particularly few studies examining whether regulatory abilities acquired training transfer to novel tasks or contexts (Loriette et al., 2021). Alternatively, the divergent effects may reflect modality-specific regulation mechanisms: the training paradigm involved explicit regulation of negative stimuli through reappraisals, whereas the cold pressor test elicited physiological pain triggering autonomic responses. Although the lateral PFC is consistently recruited across different stimulus representations of negative affect (Čeko et al., 2022), explicit and implicit emotion regulation rely on distinct but complementary processes (Gyurak et al., 2011). In the context of our study, neurofeedback may have more readily enhanced implicit regulatory pathways independent of subjective emotional awareness, rather than explicit appraisals that require deliberate processing.

### 4.4 Limitations

While the present study illustrates the success of a single-session neurofeedback training protocol in enhancing brain activity and transfer to regulation within a novel stress induction task, we consider several limitations. The durability of improvements towards stress regulation remain unclear without follow-up sessions, and findings of regulatory benefits would be further substantiated by well-validated physiological biomarkers of stress such as cortisol levels and skin conductance. In addition, though this study chiefly examined effects of neurofeedback training with a randomized-controlled design and controlled for variables such as baseline pain and self-efficacy measures, it must be acknowledged that individual differences in attention and motivation (Cohen Kadosh & Staunton, 2019) as well as intrinsic self-regulation capacity (Haugg et al., 2020; Zhao et al., 2021) may contribute to inter-individual variability in learning success. Future investigations should bear in mind factors contributing optimal neurofeedback learning to design protocols, and carefully consider implementing multiple sessions and robust assessments at different timepoints to illuminate the extent to which maintenance effects persist beyond the immediate training context.

### 4.5 Implications for future studies and approaches to global health

The distinctive utility of neurofeedback training lies in developing a personalized approach to learning regulation with real-time feedback: through individualized targeting within cortical regions of established regulatory circuitry, participants can learn and adapt regulatory strategies according to their own unique neural activation patterns. This is important given emotional reactivity is largely heterogeneous across individuals (Hamann & Canli, 2004) and influences of the selection and effective deployment of strategies (Shafir et al., 2016). For future directions, training paradigms can be further adapted to account for differences in emotional reactivity and context-dependent factors such as perceived controllability and stress intensity (Troy et al., 2013; Silvers et al., 2015). Additionally, incorporating more ecologically valid stimuli to further support transfer of regulatory skills to daily life (Zhou & Becker, 2025). Through implementing these considerations, personalized neurofeedback protocols would optimize regulatory strategies closely relevant to day-to-day challenges, ensuring that learning translates into adaptive stress regulation under ecologically valid conditions.

From an implementation perspective, fNIRS represents a promising avenue for neurofeedback interventions relative to other functional neuroimaging modalities, as its non-invasiveness, portability, and cost-effectiveness render it suitable for integration in real-world environments (Klein et al., 2024). The meaningful stress regulation benefits observed in the present study indicate potential applicability of fNIRS neurofeedback in augmenting conventional evidence-based treatments. With prior research demonstrating that supplementing cognitive behavioral therapy with neurofeedback sessions enhances therapeutic outcomes (MacDuffie et al., 2018; Compère et al., 2023), integrating fNIRS neurofeedback may complement intervention approaches to enhance stress regulatory capabilities preventatively in subclinical populations. fNIRS neurofeedback emerges at the intersection of advancing wearable systems in naturalistic settings and the development of brain-computer interfaces (Pinti et al., 2018; von Lühmann et al., 2021), uniquely positioned to integrate and harness innovations from both of these technological frontiers. Altogether, these multifaceted, converging advantages establish fNIRS neurofeedback as a promising translational pathway for stress regulation that can be scaled within existing preventative intervention frameworks to increase resilience across diverse populations and contexts.

## 5 Conclusion

The present investigation demonstrates that a single session of real-time fNIRS neurofeedback targeting the lateral PFC successfully engages specific neural learning processes, facilitating regulatory control over prefrontal activation that effectively enhances stress regulation capabilities in healthy individuals. Critically, regulation skills acquired during training transferred to novel stressful contexts, establishing that neurofeedback promotes adaptive learning of regulatory abilities that reduce stress reactivity within ecologically valid settings. This neuroscientifically-grounded approach, validated through rigorous pre-registered double-blind methodology, integrates established neural mechanisms with evidence-based psychological strategies, producing robust outcomes that extend beyond conventional cognitive training. Our findings establish fNIRS neurofeedback as a promising preventative approach for at-risk populations, offering a targeted, accessible, and scalable neuromodulatory intervention that can be integrated within existing frameworks to address the growing global burden of mental health challenges through enhanced stress resilience.

## Disclosures

The authors declare that there are no competing interests.

## Code, Data, and Materials

The data that support the findings of this article are available from the authors upon request, and with permission from the University of Hong Kong.

## Acknowledgments

The authors thank Michael Lührs for his valuable guidance and recommendations in establishing the neurofeedback protocol and system configuration. This work was supported by the STI 2030–Major Projects (Grant No.2022ZD0208500), and a start-up grant from the University of Hong Kong.

## Author Biographies

**Michelle Hei Lam Tsang** is a PhD student in the Department of Psychology at the University of Hong Kong. She received her MSc degree in cognitive neuroscience from University College London. Her research focuses on investigating cognitive and affective regulation mechanisms, with current work on neurofeedback approaches and future interests in computational methods for mental health applications.

**Judy Chuyi Chen** received her Bachelor of Social Science degree in Psychology at the University of Hong Kong. Her undergraduate thesis explored the potential applications of fNIRS neurofeedback’s applicable potential in emotion regulation. She is currently pursuing her Master’s degree with special interests in integrating neuroimaging and neurofeedback approaches to address emotion dysregulation disorders.

**Heng Jiang** is a cognitive neuroscience PhD candidate at the University of Electronic Science and Technology of China. His research focuses on emotion regulation and empathy, particularly among individuals with mental disorders, using a combination of neuroimaging, behavioral, and clinical approaches.

**Benjamin Becker** received his PhD in psychology from Heinrich Heine University Duesseldorf (Germany). Currently, he is a full professor at the University of Hong Kong, adjunct professor at the University Electronic Science and Technology in Chengdu (China), and the director of the Affective and Motivational Neuroscience Laboratory (www.Beckerlab.org). His research combines neuroimaging with pharmacological and closed-loop neuromodulation approaches to develop novel treatments for psychiatric disorders.

## Appendix: Supplemental Material

**Table 1.** Consensus on the reporting and experimental design of clinical and cognitive-behavioral neurofeedback studies (CRED-nf) checklist (Ros et al., 2020)

| Domain | Item # | Checklist item | Reported on page # |
| --- | --- | --- | --- |
| **Pre-experiment** |  |  |  |
|  | 1a | Pre-register experimental protocol and planned analyses | pp.9 |
|  | 1b | Justify sample size | pp.9 |
| **Control groups** |  |  |  |
|  | 2a | Employ control group(s) or control condition(s) | pp.10 |
|  | 2b | When leveraging experimental designs where a double-blind is possible, use a double-blind | pp.10 |
|  | 2c | Blind those who rate the outcomes, and when possible, the statisticians involved |  |
|  | 2d | Examine to what extent participants and experimenters remain blinded | pp.10 |
|  | 2e | In clinical efficacy studies, employ a standard-of-care intervention group as a benchmark for improvement |  |
| **Control measures** |  |  |  |
|  | 3a | Collect data on psychosocial factors | pp.20 |
|  | 3b | Report whether participants were provided with a strategy | pp.11 |
|  | 3c | Report the strategies participants used | pp.30 |
|  | 3d | Report methods used for online-data processing and artifact correction | pp.16 |
|  | 3e | Report condition and group effects for artifacts |  |
| **Feedback specifications** |  |  |  |
|  | 4a | Report how the online-feature extraction was defined | pp.16 |
|  | 4b | Report and justify the reinforcement schedule | pp.16 |
|  | 4c | Report the feedback modality and content | pp.16 |
|  | 4d | Collect and report all brain activity variable(s) and/or contrasts used for feedback, as displayed to experimental participants | pp.16 |
|  | 4e | Report the hardware and software used | pp.14,17 |
| **Outcome measures** |  |  |  |
| Brain | 5a | Report neurofeedback regulation success based on the feedback signal | pp.21,22 |
|  | 5b | Plot within-session and between-session regulation blocks of feedback variable(s), as well as pre-to-post resting baselines or contrasts | pp.21,22 |
|  | 5c | Statistically compare the experimental condition/group to the control condition(s)/group(s) (not only each group to baseline measures) | pp.21,22 |
| Behaviour | 6a | Include measures of clinical or behavioural significance, defined a priori, and describe whether they were reached | pp.24,25 |
|  | 6b | Run correlational analyses between regulation success and behavioural outcomes | pp.26,27 |
| **Data storage** |  |  |  |
|  | 7a | Upload all materials, analysis scripts, code, and raw data used for analyses, as well as final values, to an open access data repository, when feasible |  |

\*Darker shaded boxes represent essential checklist items; lightly shaded boxes represent encouraged checklist items.

**Table 2.** Locations of channels based on probe projection in AtlasViewer (v.2.44.0; Aasted et al., 2015).

| Channel |  | MNI coordinates |  |  | AAL region |
| --- | --- | --- | --- | --- | --- |
| Source | Detector | x | y | z |  |
| 1 | 1 | -12 | 72 | 1 | Frontal_Med_Orb_L |
| 1 | 2 | -29 | 63 | 0 | Frontal_Sup_L |
| 1 | 3 | -19 | 60 | 6 | Frontal_Sup_L |
| 2 | 2 | -40 | 59 | 10 | Frontal_Mid_L |
| 2 | 3 | -21 | 47 | 12 | Frontal_Mid_L |
| 2 | 4 | -42 | 48 | 17 | Frontal_Mid_L |
| 3 | 3 | -11 | 48 | 24 | Frontal_Sup_Medial_L |
| 3 | 5 | -9 | 44 | 32 | Frontal_Sup_Medial_L |
| 4 | 2 | -38 | 49 | 1 | Frontal_Mid_L |
| 4 | 4 | -36 | 43 | 10 | Frontal_Mid_L |
| 4 | 6 | -36 | 22 | 7 | Frontal_Inf_Tri_L |
| 5 | 3 | -19 | 46 | 21 | Frontal_Sup_L |
| 5 | 4 | -37 | 37 | 20 | Frontal_Mid_L |
| 5 | 5 | -19 | 41 | 36 | Frontal_Mid_L |
| 5 | 7 | -45 | 34 | 42 | Frontal_Mid_L |
| 6 | 4 | -35 | 22 | 22 | Frontal_Inf_Tri_L |
| 6 | 6 | -48 | 11 | 16 | Frontal_Inf_Oper_L |
| 6 | 7 | -53 | 18 | 37 | Frontal_Inf_Oper_L |
| 7 | 5 | -20 | 33 | 59 | Frontal_Sup_L |
| 7 | 7 | -37 | 18 | 59 | Frontal_Mid_L |
| 7 | 8 | -9 | 16 | 60 | Frontal_Sup_L |
| 8 | 5 | -9 | 49 | 54 | Frontal_Sup_L |
| 8 | 8 | 1 | 33 | 66 | Supp_Motor_Area_L |
| 8 | 11 | 13 | 48 | 54 | Frontal_Sup_Medial_R |
| 9 | 1 | 15 | 70 | 0 | Frontal_Med_Orb_R |
| 9 | 9 | 24 | 58 | 10 | Frontal_Sup_R |
| 9 | 10 | 36 | 66 | 1 | Frontal_Sup_Orb_R |
| 10 | 9 | 32 | 58 | 21 | Frontal_Mid_R |
| 10 | 10 | 44 | 57 | 6 | Frontal_Mid_R |
| 10 | 12 | 45 | 41 | 17 | Frontal_Inf_Tri_R |
| 11 | 9 | 22 | 46 | 23 | Frontal_Sup_R |
| 11 | 11 | 25 | 38 | 37 | Frontal_Sup_R |
| 11 | 12 | 38 | 26 | 17 | Frontal_Inf_Tri_R |
| 11 | 13 | 38 | 17 | 29 | Frontal_Inf_Oper_R |
| 12 | 10 | 55 | 52 | -1 | Frontal_Inf_Tri_R |
| 12 | 12 | 52 | 43 | 9 | Frontal_Inf_Tri_R |
| 12 | 14 | 59 | 22 | 1 | Frontal_Inf_Oper_R |
| 13 | 8 | 14 | 20 | 73 | Supp_Motor_Area_R |
| 13 | 11 | 15 | 22 | 46 | Cingulum_Mid_R |
| 13 | 13 | 37 | 14 | 55 | Frontal_Sup_R |
| 14 | 12 | 63 | 30 | 26 | Frontal_Inf_Tri_R |
| 14 | 13 | 55 | 15 | 36 | Precentral_R |
| 14 | 14 | 67 | 14 | 15 | Precentral_R |
| 15 | 9 | 16 | 52 | 27 | Frontal_Sup_R |
| 15 | 11 | 17 | 53 | 47 | Frontal_Sup_R |
| 16 | 1 | 0 | 55 | 14 | Frontal_Sup_Medial_L |
| 16 | 3 | -11 | 58 | 22 | Frontal_Sup_L |
| 16 | 9 | 15 | 60 | 24 | Frontal_Sup_R |

## Notes

### Competing Interest Statement

The authors have declared no competing interest.

### Clinical Trial

NCT06866028

### Author Declarations

Human Research Ethics Committee for Non-Clinical Faculties of the University of Hong Kong gave ethics approval (reference number: EA240016).

## References

1. Aasted, C. M., Yücel, M. A., Cooper, R. J., Dubb, J., Tsuzuki, D., Becerra, L., Petkov, M. P., Borsook, D., Dan, I., & Boas, D. A. (2015). Anatomical guidance for functional near-infrared spectroscopy: *AtlasViewer tutorial*. Neurophotonics, 2(2), 020801. 10.1117/1.NPh.2.2.020801

2. Aranyi, G., Pecune, F., Charles, F., Pelachaud, C., & Cavazza, M. (2016). Affective interaction with a virtual character through an fNIRS brain-computer interface. Frontiers in Computational Neuroscience, 10(70). 10.3389/fncom.2016.00070

3. Barker, J. W., Aarabi, A., & Huppert, T. J. (2013). Autoregressive model based algorithm for correcting motion and serially correlated errors in fNIRS. Biomedical Optics Express, 4(8), 1366–1379. 10.1364/BOE.4.001366

4. Beauregard, M., Lévesque, J., & Vincent Paquette, V. (2004). Neural basis of conscious and voluntary self-regulation of emotion. In M. Beauregard (Ed.) Consciousness, Emotional Self-Regulation and the Brain (1st ed.; pp. 174–205). John Benjamins Publishing Company. 10.1075/aicr.54

5. Beck, A. T., Steer, R. A., & Brown, G. (1996). Manual for the Beck Depression Inventory–II. Psychological Corporation.

6. Berboth, S., & Morawetz, C. (2021). Amygdala-prefrontal connectivity during emotion regulation: A meta-analysis of psychophysiological interactions. Neuropsychologia, 153, 107767. 10.1016/j.neuropsychologia.2021.107767

7. Bradley, M. M., & Lang, P. J. (1994). Measuring emotion: The Self-Assessment Manikin and the semantic differential. Journal of Behavior Therapy and Experimental Psychiatry, 25(1), 49–59. 10.1016/0005-7916(94)90063-9

8. Bradley, M. M., & Lang, P. J. (2007). The International Affective Picture System (IAPS) in the study of emotion and attention. In J. A. Coan & J. J. B. Allen (Eds.), Handbook of emotion elicitation and assessment (pp. 29–46). Oxford University Press.

9. Buhle, J. T., Silvers, J. A., Wager, T. D., Lopez, R., Onyemekwu, C., Kober, H., Weber, J., & Ochsner, K. N. (2014). Cognitive reappraisal of emotion: A meta-analysis of human neuroimaging studies. Cerebral Cortex, 24(11), 2981–90. 10.1093/cercor/bht154

10. Caspi, A., Houts, R. M., Belsky, D. W., Goldman-Mellor, S. J., Harrington, H., Israel, S., Meier, M. H., Ramrakha, S., Shalev, I., Poulton, R., & Moffitt, T. E. (2014). The p factor: One general psychopathology factor in the structure of psychiatric disorders? Clinical Psychological Science, 2(2), 119–137. 10.1177/2167702613497473

11. Čeko, M., Kragel, P. A., Woo, C. W., López-Solà, M., & Wager, T. D. (2022). Common and stimulus-type-specific brain representations of negative affect. Nature Neuroscience, 25(6), 760–770. 10.1038/s41593-022-01082-w

12. Chiasson, P., Boylan, M. R., Elhamiasl, M., Pruitt, J. M., Ranjan, S., Riels, K., Sahoo, A. K., Mirifar, A., & Keil, A. (2023). Effects of neurofeedback training on performance in laboratory tasks: A systematic review. International Journal of Psychophysiology, 189, 42–56. 10.1016/j.ijpsycho.2023.04.005

13. Cohen, S., Kamarck, T., & Mermelstein, R. (1983). A global measure of perceived stress. Journal of Health and Social Behavior, 24(4), 386–396.

14. Cohen Kadosh, K., & Staunton, G. (2019). A systematic review of the psychological factors that influence neurofeedback learning outcomes. NeuroImage, 185, 545–555. https://doi-org.eproxy.lib.hku.hk/10.1016/j.neuroimage.2018.10.021

15. Costa, P. T., & McCrae, R. R. (1992). Normal personality assessment in clinical practice: The NEO Personality Inventory. Psychological Assessment, 4(1), 5–13.

16. Compère, L., Siegle, G. J., Riley, E., Lazzaro, S., Strege, M., Pacoe, E., Canovali, G., Barb, S., Huppert, T., & Young, K. (2023). Enhanced efficacy of CBT following augmentation with amygdala rtfMRI neurofeedback in depression. Journal of Affective Disorders, 339, 495–501. 10.1016/j.jad.2023.07.063

17. COVID-19 Mental Disorders Collaborators (2021). Global prevalence and burden of depressive and anxiety disorders in 204 countries and territories in 2020 due to the COVID-19 pandemic. Lancet, 398(10312), 1700–1712. 10.1016/S0140-6736(21)02143-7

18. Crone, D. L., Bode, S., Murawski, C., & Laham, S. M. (2018). The Socio-Moral Image Database (SMID): A novel stimulus set for the study of social, moral and affective processes. PLOS One, 13(1), e0190954. 10.1371/journal.pone.0190954

19. Etkin, A., Büchel, C., & Gross, J. J. (2015). The neural bases of emotion regulation. Nature Reviews Neuroscience, 16(11), 693–700. 10.1038/nrn4044

20. Friedman, N.P., Robbins, T.W. (2022). The role of prefrontal cortex in cognitive control and executive function. Neuropsychopharmacology, 47, 72–89. 10.1038/s41386-021-01132-0

21. Gaume, A., Vialatte, A., Mora-Sánchez, A., Ramdani, C., & Vialatte, F. B. (2016). A psychoengineering paradigm for the neurocognitive mechanisms of biofeedback and neurofeedback. Neuroscience & Biobehavioral Reviews, 68, 891–910. 10.1016/j.neubiorev.2016.06.012

22. Godet, A., Serrand, Y., Léger, B., Moirand, R., Bannier, E., Val-Laillet, D., & Coquery, N. (2024). Functional near-infrared spectroscopy-based neurofeedback training targeting the dorsolateral prefrontal cortex induces changes in cortico-striatal functional connectivity. Scientific Reports, 14(1), 20025–20016. 10.1038/s41598-024-69863-w

23. Goodwin, J. R., Gaudet, C. R., & Berger, A. J. (2014). Short-channel functional near-infrared spectroscopy regressions improve when source-detector separation is reduced. Neurophotonics, 1(1), 015002. 10.1117/1.NPh.1.1.015002

24. Gross, J. J. (2002). Emotion regulation: Affective, cognitive, and social consequences. Psychophysiology, 39(3), 281–291. 10.1017/s0048577201393198

25. Gyurak, A., Gross, J. J., & Etkin, A. (2011). Explicit and implicit emotion regulation: A dual-process framework. Cognition & Emotion, 25(3), 400–412. 10.1080/02699931.2010.544160

26. Hamann, S., & Canli, T. (2004). Individual differences in emotion processing. Current Opinion in Neurobiology, 14(2), 233–238. 10.1016/j.conb.2004.03.010

27. Haugg, A., Sladky, R., Skouras, S., McDonald, A., Craddock, C., Kirschner, M., Herdener, M., Koush, Y., Papoutsi, M., Keynan, J. N., Hendler, T., Cohen Kadosh, K., Zich, C., MacInnes, J., Adcock, R. A., Dickerson, K., Chen, N. K., Young, K., Bodurka, J., … Scharnowski, F. (2020). Can we predict real-time fMRI neurofeedback learning success from pretraining brain activity? Human Brain Mapping, 41(14), 3839–3854. 10.1002/hbm.25089

28. Hou, X., Xiao, X., Gong, Y., Li, Z., Chen, A., & Zhu, C. (2021). Functional near-infrared spectroscopy neurofeedback enhances human spatial memory. Frontiers in Human Neuroscience, 15, 681193. 10.3389/fnhum.2021.681193

29. Jiang, H., He, J., Zimmermann, K., Zhou, X., Gan, X., Ferraro, S., Wang, L., Zhou, B., Li, L., Kendrick, K., Zhao, W., Yao, D., Yuan, T., Zhou, F., & Becker, B. (2025). Common and distinct neurofunctional signatures of emotion regulation strategies in dynamic naturalistic contexts and their clinical translation. medRxiv. 10.1101/2025.05.29.25328539

30. Keller, M., Zweerings, J., Klasen, M., Zvyagintsev, M., Iglesias, J., Mendoza Quiñones, R., & Mathiak, K. (2021). fMRI neurofeedback-enhanced cognitive reappraisal training in depression: A double-blind comparison of left and right vlPFC regulation. Frontiers in Psychiatry, 12, 715898–715898. 10.3389/fpsyt.2021.715898

31. Kessler, R. C., Andrews, G., Colpe, L. J., Hiripi, E., Mroczek, D. K., Normand, S. L., Walters, E. E., & Zaslavsky, A. M. (2002). Short screening scales to monitor population prevalences and trends in non-specific psychological distress. Psychological Medicine, 32(6), 959–976. 10.1017/s0033291702006074

32. Kessler, R. C., Berglund, P., Demler, O., Jin, R., Merikangas, K. R., & Walters, E. E. (2005). Lifetime prevalence and age-of-onset distributions of DSM-IV disorders in the National Comorbidity Survey Replication. Archives of General Psychiatry, 62(6), 593–602. 10.1001/archpsyc.62.6.593

33. Kieling, C., Buchweitz, C., Caye, A., Silvani, J., Ameis, S. H., Brunoni, A. R., Cost, K. T., Courtney, D. B., Georgiades, K., Merikangas, K. R., Henderson, J. L., Polanczyk, G. V., Rohde, L. A., Salum, G. A., & Szatmari, P. (2024). Worldwide prevalence and disability from mental disorders across childhood and adolescence: Evidence from the global burden of disease study. JAMA Psychiatry, 81(4), 347–356. 10.1001/jamapsychiatry.2023.5051

34. Kimmig, A. S., Dresler, T., Hudak, J., Haeussinger, F. B., Wildgruber, D., Fallgatter, A. J., Ehlis, A. C., & Kreifelts, B. (2019). Feasibility of NIRS-based neurofeedback training in social anxiety disorder: Behavioral and neural correlates. Journal of Neural Transmission, 126(9), 1175–1185. 10.1007/s00702-018-1954-5

35. Klein, F., Kohl, S. H., Lührs, M., Mehler, D. M. A., & Sorger, B. (2024). From lab to life: challenges and perspectives of fNIRS for haemodynamic-based neurofeedback in real-world environments. Philosophical Transactions of the Royal Society of London. Series B: Biological Sciences, 379(1915), 20230087. 10.1098/rstb.2023.0087

36. Krause, F., & Lindemann, O. (2014). Expyriment: A Python library for cognitive and neuroscientific experiments. Behavior Research Methods, 46(2), 416–428. 10.3758/s13428-013-0390-6

37. Krause, F., Linden, D. E. J., & Hermans, E. J. (2024). Getting stress-related disorders under control: The untapped potential of neurofeedback. Trends in Neurosciences, 47(10), 766–776. 10.1016/j.tins.2024.08.007

38. Kohl, S. H., Mehler, D. M. A., Lührs, M., Thibault, R. T., Konrad, K., & Sorger, B. (2020). The potential of functional near-infrared spectroscopy-based neurofeedback—A systematic review and recommendations for best practice. Frontiers in Neuroscience, 14, 594–594. 10.3389/fnins.2020.00594

39. Kohn, N., Eickhoff, S. B., Scheller, M., Laird, A. R., Fox, P. T., & Habel, U. (2014). Neural network of cognitive emotion regulation--an ALE meta-analysis and MACM analysis. NeuroImage, 87, 345–355. 10.1016/j.neuroimage.2013.11.001

40. Li, K., Jiang, Y., Gong, Y., Zhao, W., Zhao, Z., Liu, X., Kendrick, K. M., Zhu, C., & Becker, B. (2019). Functional near-infrared spectroscopy-informed neurofeedback: regional-specific modulation of lateral orbitofrontal activation and cognitive flexibility. Neurophotonics, 6(2), 025011. 10.1117/1.NPh.6.2.025011

41. Li, K., Yang, J., Becker, B., & Li, X. (2023). Functional near-infrared spectroscopy neurofeedback of dorsolateral prefrontal cortex enhances human spatial working memory. Neurophotonics, 10(2), 025011. 10.1117/1.NPh.10.2.025011

42. Liu, Y., Ren, Y., Liu, C., Chen, X., Li, D., Peng, J., Tan, L., & Ma, Q. (2025). Global burden of mental disorders in children and adolescents before and during the COVID-19 pandemic: Evidence from the Global Burden of Disease Study 2021. Psychological Medicine, 55, e90. 10.1017/S0033291725000649

43. Linhartová, P., Látalová, A., Kóša, B., Kašpárek, T., Schmahl, C., & Paret, C. (2019). fMRI neurofeedback in emotion regulation: A literature review. NeuroImage, 193, 75–92. 10.1016/j.neuroimage.2019.03.011

44. Loriette, C., Ziane, C., & Ben Hamed, S. (2021). Neurofeedback for cognitive enhancement and intervention and brain plasticity. Revue Neurologique, 177(9), 1133–1144. 10.1016/j.neurol.2021.08.004

45. Lubianiker, N., Goldway, N., Fruchtman-Steinbok, T., Paret, C., Keynan, J. N., Singer, N., Cohen, A., Kadosh, K. C., Linden, D. E. J., & Hendler, T. (2019). Process-based framework for precise neuromodulation. Nature Human Behaviour, 3(5), 436–445. 10.1038/s41562-019-0573-y

46. Lührs, M., & Goebel, R. (2017). Turbo-Satori: a neurofeedback and brain-computer interface toolbox for real-time functional near-infrared spectroscopy (Version 2.2.2) [Computer software]. Neurophotonics, 4(4), 041504. 10.1117/1.NPh.4.4.041504

47. MacDuffie, K. E., MacInnes, J., Dickerson, K. C., Eddington, K. M., Strauman, T. J., & Adcock, R. A. (2018). Single session real-time fMRI neurofeedback has a lasting impact on cognitive behavioral therapy strategies. NeuroImage. Clinical, 19, 868–875. 10.1016/j.nicl.2018.06.009

48. Marchewka, A., Zurawski, Ł., Jednoróg, K., & Grabowska, A. (2014). The Nencki Affective Picture System (NAPS): Introduction to a novel, standardized, wide-range, high-quality, realistic picture database. Behavior Research Methods, 46(2), 596–610. 10.3758/s13428-013-0379-1

49. Mihov, Y., Kendrick, K. M., Becker, B., Zschernack, J., Reich, H., Maier, W., Keysers, C., & Hurlemann, R. (2013). Mirroring fear in the absence of a functional amygdala. Biological Psychiatry, 73(7), e9–e11. 10.1016/j.biopsych.2012.10.029

50. Morawetz, C., Bode, S., Derntl, B., & Heekeren, H. R. (2017). The effect of strategies, goals and stimulus material on the neural mechanisms of emotion regulation: A meta-analysis of fMRI studies. Neuroscience and Biobehavioral Reviews, 72, 111–128. 10.1016/j.neubiorev.2016.11.014

51. Ochsner, K. N., Silvers, J. A., & Buhle, J. T. (2012). Functional imaging studies of emotion regulation: a synthetic review and evolving model of the cognitive control of emotion. Annals of the New York Academy of Sciences, 1251, E1–E24. 10.1111/j.1749-6632.2012.06751.x

52. Paret, C., Goldway, N., Zich, C., Keynan, J. N., Hendler, T., Linden, D., & Cohen Kadosh, K. (2019). Current progress in real-time functional magnetic resonance-based neurofeedback: Methodological challenges and achievements. NeuroImage, 202, 116107. 10.1016/j.neuroimage.2019.116107

53. Paret, C., Kluetsch, R., Zaehringer, J., Ruf, M., Demirakca, T., Bohus, M., Ende, G., & Schmahl, C. (2016). Alterations of amygdala-prefrontal connectivity with real-time fMRI neurofeedback in BPD patients. Social Cognitive and Affective Neuroscience, 11(6), 952–960. 10.1093/scan/nsw016

54. Peirce, J., Gray, J. R., Simpson, S., MacAskill, M., Höchenberger, R., Sogo, H., Kastman, E., & Lindeløv, J. K. (2019). *PsychoPy2: Experiments in behavior made easy* (Version 2024.1.4) [Computer software]. Behavior Research Methods, 51(1), 195–203. 10.3758/s13428-018-01193-y

55. Picó-Pérez, M., Radua, J., Steward, T., Menchón, J. M., & Soriano-Mas, C. (2017). Emotion regulation in mood and anxiety disorders: A meta-analysis of fMRI cognitive reappraisal studies. Progress in Neuro-Psychopharmacology & Biological Psychiatry, 79(Pt B), 96–104. 10.1016/j.pnpbp.2017.06.001

56. Pinti, P., Aichelburg, C., Gilbert, S., Hamilton, A., Hirsch, J., Burgess, P., & Tachtsidis, I. (2018). A review on the use of wearable functional near-infrared spectroscopy in naturalistic environments. Japanese Psychological Research, 60(4), 347–373. 10.1111/jpr.12206

57. Ray, R. D., & Zald, D. H. (2012). Anatomical insights into the interaction of emotion and cognition in the prefrontal cortex. Neuroscience and Biobehavioral Reviews, 36(1), 479–501. 10.1016/j.neubiorev.2011.08.005

58. Ros, T., Enriquez-Geppert, S., Zotev, V., Young, K. D., Wood, G., Whitfield-Gabrieli, S., Wan, F., Vuilleumier, P., Vialatte, F., Van De Ville, D., Todder, D., Surmeli, T., Sulzer, J. S., Strehl, U., Sterman, M. B., Steiner, N. J., Sorger, B., Soekadar, S. R., Sitaram, R., … Thibault, R. T. (2020). Consensus on the reporting and experimental design of clinical and cognitive-behavioural neurofeedback studies (CRED-nf checklist). Brain, 143(6), 1674–1685. 10.1093/brain/awaa009

59. Ruscheweyh, R., Marziniak, M., Stumpenhorst, F., Reinholz, J., & Knecht, S. (2009). Pain sensitivity can be assessed by self-rating: Development and validation of the Pain Sensitivity Questionnaire. Pain, 146(1), 65–74. 10.1016/j.pain.2009.06.020

60. Santosa, H., Zhai, X., Fishburn, F., & Huppert, T. (2018). The NIRS Brain AnalyzIR Toolbox. Algorithms, 11(5), 73. 10.3390/a11050073

61. Santosa, H., Zhai, X., Fishburn, F., Sparto, P. J., & Huppert, T. J. (2020). Quantitative comparison of correction techniques for removing systemic physiological signal in functional near-infrared spectroscopy studies. Neurophotonics, 7(3), 035009. 10.1117/1.NPh.7.3.035009

62. Schwabe, L., Haddad, L., & Schachinger, H. (2008). HPA axis activation by a socially evaluated cold-pressor test. Psychoneuroendocrinology, 33(6), 890–895. 10.1016/j.psyneuen.2008.03.001

63. Schwabe, L., & Schächinger, H. (2018). Ten years of research with the Socially Evaluated Cold Pressor Test: Data from the past and guidelines for the future. Psychoneuroendocrinology, 92, 155–161. 10.1016/j.psyneuen.2018.03.010

64. Shafir, R., Thiruchselvam, R., Suri, G., Gross, J. J., & Sheppes, G. (2016). Neural processing of emotional-intensity predicts emotion regulation choice. Social Cognitive and Affective Neuroscience, 11(12), 1863–1871. 10.1093/scan/nsw114

65. Sherer, M., Maddux, J. E., Mercandante, B., Prentice-Dunn, S., Jacobs, B., & Rogers, R. W. (1982). The self-efficacy scale: Construction and validation. Psychological Reports, 51(2), 663–671.

66. Silvers, J. A., Weber, J., Wager, T. D., & Ochsner, K. N. (2015). Bad and worse: neural systems underlying reappraisal of high-and low-intensity negative emotions. Social Cognitive and Affective Neuroscience, 10(2), 172–179. 10.1093/scan/nsu043

67. Sitaram, R., Ros, T., Stoeckel, L., Haller, S., Scharnowski, F., Lewis-Peacock, J., Weiskopf, N., Blefari, M. L., Rana, M., Oblak, E., Birbaumer, N., & Sulzer, J. (2017). Closed-loop brain training: the science of neurofeedback. Nature Reviews Neuroscience, 18(2), 86–100. 10.1038/nrn.2016.164

68. Soekadar, S. R., Kohl, S. H., Mihara, M., & von Lühmann, A. (2021). Optical brain imaging and its application to neurofeedback. NeuroImage: Clinical, 30, 102577. 10.1016/j.nicl.2021.102577

69. Solmi, M., Radua, J., Olivola, M., Croce, E., Soardo, L., Salazar de Pablo, G., Il Shin, J., Kirkbride, J. B., Jones, P., Kim, J. H., Kim, J. Y., Carvalho, A. F., Seeman, M. V., Correll, C. U., & Fusar-Poli, P. (2022). Age at onset of mental disorders worldwide: Large-scale meta-analysis of 192 epidemiological studies. Molecular Psychiatry, 27(1), 281–295. 10.1038/s41380-021-01161-7

70. Steinbrink, J., Villringer, A., Kempf, F., Haux, D., Boden, S., & Obrig, H. (2006). Illuminating the BOLD signal: Combined fMRI-fNIRS studies. Magnetic Resonance Imaging, 24(4), 495–505. 10.1016/j.mri.2005.12.034

71. Stoeckel, L. E., Garrison, K. A., Ghosh, S., Wighton, P., Hanlon, C. A., Gilman, J. M., Greer, S., Turk-Browne, N. B., deBettencourt, M. T., Scheinost, D., Craddock, C., Thompson, T., Calderon, V., Bauer, C. C., George, M., Breiter, H. C., Whitfield-Gabrieli, S., Gabrieli, J. D., LaConte, S. M., … Evins, A. E. (2014). Optimizing real time fMRI neurofeedback for therapeutic discovery and development. NeuroImage. Clinical, 5, 245–255. 10.1016/j.nicl.2014.07.002

72. Spielberger, C., Gorsuch, R., Lushene, R., Vagg, P., & Jacobs, G. (1983). Manual for the State-Trait Anxiety Inventory. Consulting Psychologists Press.

73. Troy, A. S., Shallcross, A. J., & Mauss, I. B. (2013). A person-by-situation approach to emotion regulation: Cognitive reappraisal can either help or hurt, depending on the context. Psychological Science, 24(12), 2505–2514. 10.1177/0956797613496434

74. Tang, Y., Chen, Z., Jiang, Y., Zhu, C., & Chen, A. (2021). From reversal to normal: Robust improvement in conflict adaptation through real-time functional near infrared spectroscopy-based neurofeedback training. Neuropsychologia, 157, 107866. 10.1016/j.neuropsychologia.2021.107866

75. Thibault, R. T., MacPherson, A., Lifshitz, M., Roth, R. R., & Raz, A. (2018). Neurofeedback with fMRI: A critical systematic review. NeuroImage, 172, 786–807. 10.1016/j.neuroimage.2017.12.071

76. Trambaiolli, L. R., Biazoli, C. E., Jr, Cravo, A. M., Falk, T. H., & Sato, J. R. (2018). Functional near-infrared spectroscopy-based affective neurofeedback: Feedback effect, illiteracy phenomena, and whole-connectivity profiles. Neurophotonics, 5(3), 035009. 10.1117/1.NPh.5.3.035009

77. Tupak, S. V., Dresler, T., Guhn, A., Ehlis, A. C., Fallgatter, A. J., Pauli, P., & Herrmann, M. J. (2014). Implicit emotion regulation in the presence of threat: Neural and autonomic correlates. NeuroImage, 85(1), 372–379. 10.1016/j.neuroimage.2013.09.066

78. Uhlhaas, P. J., Davey, C. G., Mehta, U. M., Shah, J., Torous, J., Allen, N. B., Avenevoli, S., Bella-Awusah, T., Chanen, A., Chen, E. Y. H., Correll, C. U., Do, K. Q., Fisher, H. L., Frangou, S., Hickie, I. B., Keshavan, M. S., Konrad, K., Lee, F. S., Liu, C. H., … Wood, S. J. (2023). Towards a youth mental health paradigm: A perspective and roadmap. Molecular Psychiatry, 28(8), 3171–3181. 10.1038/s41380-023-02202-z

79. von Lühmann, A., Zheng, Y., Ortega-Martinez, A., Kiran, S., Somers, D. C., Cronin-Golomb, A., Awad, L. N., Ellis, T. D., Boas, D. A., & Yücel, M. A. (2021). Towards Neuroscience of the Everyday World (NEW) using functional near-infrared spectroscopy. Current Opinion in Biomedical Engineering, 18, 100272. 10.1016/j.cobme.2021.100272

80. Webb, T. L., Miles, E., & Sheeran, P. (2012). Dealing with feeling: a meta-analysis of the effectiveness of strategies derived from the process model of emotion regulation. Psychological Bulletin, 138(4), 775–808. 10.1037/a0027600

81. Weierich, M. R., Kleshchova, O., Rieder, J. K., & Reilly, D. M. (2019). The Complex Affective Scene Set (COMPASS): Solving the social content problem in affective visual stimulus sets. Collabra: Psychology, 5(1), 53. 10.1525/collabra.256

82. Yang, X., Zeng, Y., Jiao, G., Gan, X., Linden, D., Hernaus, D., Zhu, C., Li, K., Yao, D., Yao, S., Jiang, Y., & Becker, B. (2024). A brief real-time fNIRS-informed neurofeedback training of the prefrontal cortex changes brain activity and connectivity during subsequent working memory challenge. Progress in Neuro-Psychopharmacology & Biological Psychiatry, 132, 110968. 10.1016/j.pnpbp.2024.110968

83. Yao, S., Becker, B., Geng, Y., Zhao, Z., Xu, X., Zhao, W., Ren, P., & Kendrick, K.M. (2016). Voluntary control of anterior insula and its functional connections is feedback-independent and increases pain empathy. NeuroImage, 130, 230–240. 10.1016/j.neuroimage.2016.02.035

84. Young, K. D., Zotev, V., Phillips, R., Misaki, M., Yuan, H., Drevets, W. C., & Bodurka, J. (2014). Real-time fMRI neurofeedback training of amygdala activity in patients with major depressive disorder. PLOS One, 9(2), e88785. 10.1371/journal.pone.0088785

85. Yu, L., Long, Q., Tang, Y., Yin, S., Chen, Z., Zhu, C., & Chen, A. (2021). Improving emotion regulation through real-time neurofeedback training on the right dorsolateral prefrontal cortex: Evidence from behavioral and brain network analyses. Frontiers in Human Neuroscience, 15, 620342. 10.3389/fnhum.2021.620342

86. Zhao, Z., Yao, S., Li, K., Sindermann, C., Zhou, F., Zhao, W., Li, J., Lührs, M., Goebel, R., Kendrick, K. M., & Becker, B. (2019). Real-time functional connectivity-informed neurofeedback of amygdala-frontal pathways reduces anxiety. Psychotherapy and Psychosomatics, 88(1), 5–15. 10.1159/000496057

87. Zhao, Z., Yao, S., Zweerings, J., Zhou, X., Zhou, F., Kendrick, K. M., Chen, H., Mathiak, K., & Becker, B. (2021). Putamen volume predicts real-time fMRI neurofeedback learning success across paradigms and neurofeedback target regions. Human Brain Mapping, 42(6), 1879–1887. 10.1002/hbm.25336

88. Zhou, F., & Becker, B. (2025). Understanding human brain function in real-world environments. PLOS Biology, 23(6), e3003210. 10.1371/journal.pbio.3003210

89. Zhang, Y., Zhang, Q., Wang, J., Zhou, M., Qing, Y., Zou, H., Li, J., Yang, C., Becker, B., Kendrick, K. M., & Yao, S. (2023). “Listen to your heart”: A novel interoceptive strategy for real-time fMRI neurofeedback training of anterior insula activity. NeuroImage, 284, 120455. 10.1016/j.neuroimage.2023.120455

90. Zhuang, Q., Xu, L., Zhou, F., Yao, S., Zheng, X., Zhou, X., Li, J., Xu, X., Fu, M., Li, K., Vatansever, D., Kendrick, K. M., & Becker, B. (2021). Segregating domain-general from emotional context-specific inhibitory control systems - ventral striatum and orbitofrontal cortex serve as emotion-cognition integration hubs. NeuroImage, 238, 118269. 10.1016/j.neuroimage.2021.118269c

91. Zilverstand, A., Parvaz, M. A., & Goldstein, R. Z. (2017). Neuroimaging cognitive reappraisal in clinical populations to define neural targets for enhancing emotion regulation. A systematic review. NeuroImage, 151, 105–116. 10.1016/j.neuroimage.2016.06.009

92. Zilverstand, A., Sorger, B., Sarkheil, P., & Goebel, R. (2015). fMRI neurofeedback facilitates anxiety regulation in females with spider phobia. Frontiers in Behavioral Neuroscience, 9, 148–148. 10.3389/fnbeh.2015.00148

93. Zimeo Morais, G. A., Balardin, J. B., & Sato, J. R. (2018). fNIRS Optodes’ Location Decider (fOLD): a toolbox for probe arrangement guided by brain regions-of-interest. Scientific Reports, 8(1), 3341. 10.1038/s41598-018-21716-z

94. Zimmermann, K., Walz, C., Derckx, R. T., Kendrick, K. M., Weber, B., Dore, B., Ochsner, K. N., Hurlemann, R., & Becker, B. (2017). Emotion regulation deficits in regular marijuana users. Human Brain Mapping, 38(8), 4270–4279. 10.1002/hbm.23671

95. Zotev, V., Krueger, F., Phillips, R., Alvarez, R. P., Simmons, W. K., Bellgowan, P., Drevets, W. C., & Bodurka, J. (2011). Self-regulation of amygdala activation using real-time fMRI neurofeedback. PLOS One, 6(9), e24522. 10.1371/journal.pone.0024522

96. Zweerings, J., Sarkheil, P., Keller, M., Dyck, M., Klasen, M., Becker, B., Gaebler, A. J., Ibrahim, C. N., Turetsky, B. I., Zvyagintsev, M., Flatten, G., & Mathiak, K. (2020). Rt-fMRI neurofeedback-guided cognitive reappraisal training modulates amygdala responsivity in posttraumatic stress disorder. NeuroImage: Clinical, 28, 102483. 10.1016/j.nicl.2020.102483

